# Heterogeneous Treatment Effect for Targeted Temperature Management After Cardiac Arrest: A Causal Machine Learning Analysis

**DOI:** 10.64898/2026.05.04.26352388

**Authors:** Michel B. Raskin, Isalis Karhu-Leperd, Carl Harris, Romain Pirracchio, Jean-Baptiste Lascarrou, Robert D. Stevens

**Affiliations:** Department of Applied Mathematics and Statistics, Whiting School of Engineering, Johns Hopkins University, Baltimore, MD, USA; Laboratory of Computational Intensive Care Medicine, Johns Hopkins University School of Medicine, Baltimore, MD, USA; School of Life Sciences, Federal Institute of Technology Lausanne (EPFL), Lausanne, Switzerland; Department of Biomedical Engineering, Whiting School of Engineering, Johns Hopkins University, Baltimore, MD, USA; Department of Anesthesia and Perioperative Care, University of California San Francisco, San Francisco, CA, USA; Médecine Intensive Réanimation, Nantes University Hospital Hôtel-Dieu, Nantes, France; Department of Anesthesiology and Critical Care Medicine, Johns Hopkins University School of Medicine, Baltimore, MD, USA

**Keywords:** cardiac arrest, targeted temperature management, therapeutic hypothermia, heterogeneous treatment effects, causal machine learning, conditional average treatment effect

## Abstract

**Objectives:** To determine whether heterogeneous treatment effects (HTE) explain the inconclusive results of targeted temperature management (TTM) trials after cardiac arrest, using causal machine learning across four datasets.

**Design:** Secondary analysis of one multicenter RCT and three observational ICU cohorts using S-learner and forest-based R-learner models to estimate conditional average treatment effects (CATE).

**Setting:** Twenty-six French ICUs (HYPERION), approximately 200 U.S. ICUs (eICU-CRD), Johns Hopkins Hospital (PMAP), and Beth Israel Deaconess Medical Center (MIMIC-IV).

**Patients:** Adults (≥18 years) with cardiac arrest; 4,507 patients across the four datasets, of whom 1,814 (40.2%) received TTM.

**Interventions:** TTM as administered clinically or per HYPERION protocol. Ascertainment: randomization (HYPERION), treatment documentation (eICU-CRD), sustained hypothermia <36°C for >12 hours (PMAP), or documented cooling device use ≥12 hours (MIMIC-IV).

**Measurements and Main Results:** The primary outcome was hospital mortality; the secondary outcome was favorable neurologic function (Cerebral Performance Category 1–2 at 90 days for HYPERION; last motor Glasgow Coma Scale = 6 for observational cohorts). Three S-learner models (XGBoost, neural network, Bayesian Additive Regression Trees) and one forest-based R-learner (CausalForestDML) estimated CATE. HTE was assessed by likelihood-ratio tests for CATE×treatment interaction, CausalForestDML 95% confidence intervals, Group Average Treatment Effects (GATES) across CATE quintiles, and SHAP feature importance. S-learner discrimination was adequate (AUROC 0.72–0.82). No model showed a significant CATE×TTM interaction in any dataset (all p > 0.05). Individual CATE confidence intervals uniformly crossed zero, and GATES showed no monotonic gradient of benefit across quintiles in any dataset.

**Conclusions:** Across four diverse datasets and multiple causal machine-learning approaches, we found no evidence of heterogeneous treatment effects for TTM after cardiac arrest. The inconclusive findings of TTM trials are unlikely explained by differential effects in identifiable subgroups defined by routinely available clinical features.

**KEY POINTS:** **Question:** Do identifiable patient subgroups derive differential benefit from targeted temperature management (TTM) after cardiac arrest?

**Findings:** In a causal machine-learning analysis of 4,507 patients across one randomized trial and three observational ICU cohorts, no model detected significant heterogeneous TTM effects on mortality or neurologic outcome.

**Meaning:** Conflicting TTM trial results are unlikely explained by differential effects in identifiable subgroups, weakening the rationale for personalized TTM strategies based on routinely available clinical features.

## INTRODUCTION

Targeted temperature management (TTM) with mild hypothermia is a standard intervention for comatose survivors of cardiac arrest following return of spontaneous circulation.^1^ The original 2002 trials demonstrated improved neurological recovery and reduced mortality, prompting widespread adoption.^1^ However, the TTM1 trial (2013) found no difference between 33°C and 36°C,^2^ and TTM2 (2021), the largest prospective trial, found no benefit of hypothermia over normothermia.^3^ The HYPERION trial (2019) showed that TTM improved neurological outcomes at 90 days in patients with non-shockable rhythms,^4^ but a subsequent individual patient data meta-analysis combining TTM1 and TTM2 (n = 2,800) found no benefit in any pre-specified subgroup.^5^ Recent analyses have further explored whether specific subgroups—including patients without bystander CPR and those with particular initial rhythms—may respond differentially to temperature management,^23–26^ motivating rigorous examination of HTE in this population.

A recurrent hypothesis is that these conflicting findings reflect heterogeneous treatment effects (HTE): certain patient subgroups benefit while others do not, and this differential response may be masked when average treatment effects are estimated.^6,7^ Conventional subgroup analyses suffer from well-known limitations: they increase Type I error through multiple comparisons, assume linear covariate–outcome relationships, and are constrained by pre-specified subgroup definitions that may miss complex interaction patterns.^6^

Causal machine learning methods address these limitations by modeling high-dimensional, non-linear interactions between covariates and treatment effects to estimate conditional average treatment effects (CATE).^8,9^ Among these, metalearners—including S-learners, T-learners, X-learners, and R-learners—have been validated in recent critical care studies.^10–12^ For example, Buell et al. used causal machine learning to identify HTE for oxygen targets in mechanically ventilated patients,^10^ and Blette et al. demonstrated heterogeneous responses to dexamethasone in patients with COVID-19.^12^

In this study, we applied S-learner and forest-based R-learner models across one randomized trial and three observational intensive care unit (ICU) datasets to determine (1) whether HTE exists for TTM after cardiac arrest, and (2) which patient characteristics, if any, are associated with differential treatment benefit.

## MATERIALS AND METHODS

### Data Sources

The study was conducted using deidentified data; institutional review board (IRB) approval and informed consent details are provided in eTable 1 (Supplemental Digital Content 1). We analyzed data from four sources, summarized in eTable 1.

*HYPERION*.^4^ A multicenter randomized controlled trial of TTM in cardiac-arrest patients with non-shockable rhythms (n = 581). As a randomized trial, HYPERION satisfies the conditional ignorability assumption by design, providing the strongest basis for causal inference in this analysis.

*eICU-CRD*.^13^ A multicenter U.S. ICU database. We identified 2,082 cardiac-arrest patients with at least two motor Glasgow Coma Scale (mGCS) recordings and a first mGCS < 6. *PMAP*. Johns Hopkins University electronic health record data. We identified 1,292 eligible patients using admission type, care location, and mGCS criteria.

*MIMIC-IV*.^14^ We identified 552 eligible patients using combined International Classification of Diseases code, chief complaint, and arrest documentation criteria.

### Patient Selection and TTM Identification

Patients were included if they were ≥18 years of age, had a cardiac arrest during the index hospitalization, and had at least two mGCS recordings. Inclusion and exclusion criteria for the non-RCT datasets are depicted in the patient-selection flowchart (eFigure 6).

TTM ascertainment was dataset-specific. In HYPERION, TTM status was determined by randomization. In eICU-CRD, TTM was identified through explicit treatment documentation. In PMAP, TTM was inferred from sustained hypothermia (<36°C for >12 consecutive hours). In MIMIC-IV, TTM was defined by documented cooling device use for ≥12 consecutive hours. TTM prevalence ranged from 30% to 50% across datasets (Table 1). Temperature trajectories by TTM group across all four datasets are shown in Figure 4. The duration, target temperature, and cooling-device type varied across datasets, which may have contributed to the variability in CATE estimates across cohorts.

**Table 1.** Population characteristics across the four study datasets. HYPERION enrolled exclusively non-shockable rhythm patients; rhythm distribution in observational cohorts is reported in eTables 16–19. Favorable neurologic = 90-day Cerebral Performance Category 1–2 (HYPERION) or last recorded motor Glasgow Coma Scale = 6 at discharge (eICU-CRD, PMAP, MIMIC-IV).

| Characteristic | HYPERION | PMAP | eICU-CRD | MIMIC-IV |
| --- | --- | --- | --- | --- |
| Patients, n | 581 | 1,292 | 2,082 | 552 |
| Age, mean (years) | 65.3 | 61.6 | 62.6 | 61.8 |
| Male sex, % | 64.2 | 57.2 | 57.6 | 62.3 |
| Race/ethnicity, % |  |  |  |  |
| White | — | 50.7 | 70.4 | 42.9 |
| Black | — | 34.5 | 16.8 | 12.5 |
| Hispanic/Latino | — | 6.1 | 3.2 | 3.1 |
| Asian | — | 0.2 | 1.1 | 3.4 |
| Native American | — | — | 1.0 | — |
| Other/Unknown | — | 8.5 | 7.5 | 38.1 |
| TTM (hypothermia), n | 284 | 621 | 631 | 278 |
| Survival, TTM, n/N | 52/284 | 168/621 | 249/631 | 110/278 |
| Survival, no TTM, n/N | 50/297 | 309/671 | 697/1,451 | 109/274 |
| Favorable neurologic, TTM, n/N | 29/284 | 102/621 | 168/631 | 91/278 |
| Favorable neurologic, no TTM, n/N | 17/297 | 214/671 | 494/1,451 | 100/274 |

### Predictive Features, Outcomes, and Missingness

Features were extracted from the first 6 hours after arrest or admission, using the first recorded measurement to avoid overlap with TTM initiation. Feature selection in the observational datasets was guided by variables available in the HYPERION trial, yielding 46–310 features per dataset (eTables 16–19).

The primary outcome was hospital mortality, applied consistently across all datasets (in-hospital mortality for eICU-CRD, PMAP, and MIMIC-IV; HYPERION in-hospital mortality was harmonized to this endpoint despite the trial’s original 90-day follow-up design). Neurological function—Cerebral Performance Category 1–2 at 90 days in HYPERION and last mGCS = 6 at discharge in the observational cohorts—was a pre-specified secondary outcome.

Missingness rates per variable and per dataset were recorded (eTables 16–19). We tested whether missingness differed between TTM and non-TTM groups using chi-square or Fisher exact tests.

### Causal Machine-Learning Framework

The assumed causal structure relating pre-arrest factors, arrest characteristics, post-ROSC severity, TTM, and outcomes is shown in a directed acyclic graph (Figure 1). Guided by this framework, we used S-learners and CausalForestDML to estimate CATE. We selected these methods because they provide complementary strengths: S-learners allow direct evaluation of predictive performance, while CausalForestDML is better suited for treatment-effect estimation in observational data through cross-fitted nuisance models and Neyman-orthogonal estimation.^9^ T-learners were not used because treatment imbalance in several cohorts would leave the smaller arm underpowered. X-learners, although specifically designed for asymmetric treatment settings,^8^ were omitted to keep the comparison between predictive (S-learner) and orthogonalized (CausalForestDML) estimators interpretable; extension to X-learner-based estimation is a direction for future work. eTable 2 summarizes the broader metalearner framework.

**Figure 1.**
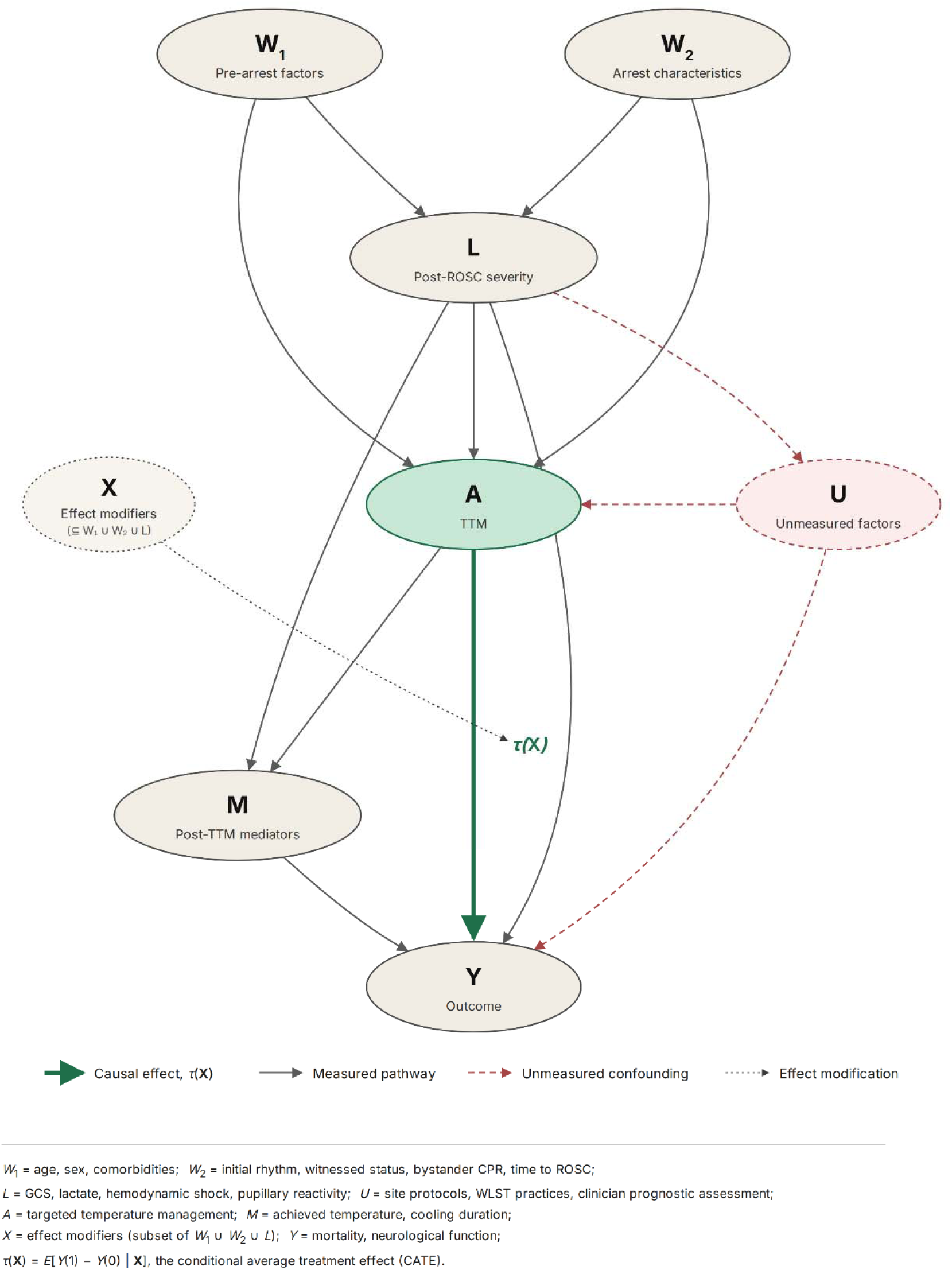
Directed acyclic graph (DAG) describing the assumed causal pathway. Pre-arrest factors and arrest characteristics influence post-ROSC severity. Post-ROSC severity either directly determines the outcome (combined with unmeasured factors) or is influenced by TTM, with post-TTM mediators on the path to outcome. ROSC = return of spontaneous circulation; TTM = targeted temperature management.

Three S-learner base models were used: XGBoost,^15^ neural networks, and Bayesian Additive Regression Trees (BART).^16^ CausalForestDML^17^ was used as a forest-based R-learner with cross-fitted nuisance models. Preprocessing included standard scaling for numerical features, one-hot encoding for categorical features, and k-nearest-neighbor imputation for missing values, as illustrated in eFigure 1.

Models were evaluated on held-out test sets. For each observational dataset, propensity scores were extracted from the CausalForestDML internal estimates. Overlap between TTM and non-TTM groups was assessed visually via propensity density plots and quantitatively by flagging scores < 0.05 or > 0.95 as potential positivity violations. Fewer than 0.3% of patients in any dataset had extreme propensity scores, suggesting adequate covariate overlap for causal estimation (eFigure 2).

### Heterogeneous Treatment Effect Assessment

HTE was assessed using four complementary approaches. First, a likelihood-ratio (LR) test compared logistic regression models with and without a CATE×TTM interaction term; a significant interaction would support HTE.^10^ Second, 95% confidence intervals for individual CATE estimates from CausalForestDML were examined. Third, Group Average Treatment Effects (GATES)^22^ were estimated by ranking patients into quintiles of predicted CATE from CausalForestDML; a joint F-test assessed whether treatment effects differed across quintiles, and Spearman rank correlation assessed monotonicity of the observed pattern (Figure 3, eTable 15). Fourth, SHAP values^21^ were used to describe the predictive importance of TTM within each S-learner model.

To evaluate the predicted probabilities used in CATE estimation, S-learner models were assessed on held-out test sets using Brier scores and calibration slopes. Calibration plots were generated for both hospital mortality (eFigure 4, eTable 4) and neurological outcome (eFigure 3, eTable 3) across all datasets.

### Ethical Statements

MIMIC-IV and eICU-CRD are publicly available via PhysioNet; their use is exempt from IRB approval. HYPERION was conducted with informed consent under its original protocol ethics approval (ClinicalTrials.gov NCT02057835).

PMAP data access and analysis were approved by the Johns Hopkins Medicine Institutional Review Board with a waiver of informed consent for retrospective deidentified data.

### Reporting Guidelines

This study adheres to the STROBE checklist for observational studies (eTable 20). The HYPERION trial component follows the CONSORT statement.^27^

## RESULTS

### Study Population

We analyzed data on 4,507 patients across four datasets. Statistically significant differences in missingness were observed for some variables (eTables 16–19). Among those that did, the majority were laboratory tests with lower missingness rates in the TTM group, which is expected given that patients receiving active temperature management typically undergo more intensive monitoring. No major clinical predictors showed significant differential missingness between groups. Within this aggregate sample, 1,814 (40.2%) received TTM. Survival with TTM ranged from 18% (HYPERION) to 40% (MIMIC-IV).

### Model Performance

All S-learner models achieved adequate discrimination on held-out test sets for the primary outcome of hospital mortality (AUROC range 0.72–0.82; Table 2). These results are consistent with prior cardiac-arrest outcome-prediction models using early clinical features.^18^

**Table 2.** Model performance and likelihood-ratio interaction-test results for hospital mortality (primary outcome) and neurological function (secondary outcome). LR p-values test the CATE×TTM interaction. AUROC and 95% CIs are from held-out test sets for S-learner models; CausalForestDML does not produce AUROC. AUROC = area under the receiver operating characteristic curve; BART = Bayesian Additive Regression Trees; CATE = conditional average treatment effect; CI = confidence interval; CPC = Cerebral Performance Category; LR = likelihood ratio; mGCS = motor Glasgow Coma Scale; TTM = targeted temperature management.

| Dataset | Outcome | Model | LR p-value | AUROC | AUROC 95% CI |
| --- | --- | --- | --- | --- | --- |
| eICU-CRD | Hospital mortality | Neural network | 0.90 | 0.75 | [0.71, 0.79] |
| eICU-CRD | Last mGCS = 6 | Neural network | 0.84 | 0.77 | [0.72, 0.80] |
| eICU-CRD | Hospital mortality | XGBoost | 0.38 | 0.74 | [0.70, 0.78] |
| eICU-CRD | Last mGCS = 6 | XGBoost | No effect | 0.74 | [0.70, 0.78] |
| eICU-CRD | Hospital mortality | BART | 0.37 | 0.77 | [0.73, 0.81] |
| eICU-CRD | Last mGCS = 6 | BART | 0.93 | 0.78 | [0.74, 0.82] |
| eICU-CRD | Hospital mortality | CausalForestDML | 0.15 | NA | NA |
| eICU-CRD | Last mGCS = 6 | CausalForestDML | 0.10 | NA | NA |
| HYPERION | Hospital mortality | Neural network | 0.23 | 0.80 | [0.71, 0.89] |
| HYPERION | 90-day CPC 1–2 | Neural network | 0.39 | 0.79 | [0.76, 0.86] |
| HYPERION | Hospital mortality | XGBoost | No effect | 0.82 | [0.73, 0.90] |
| HYPERION | 90-day CPC 1–2 | XGBoost | No effect | 0.81 | [0.68, 0.91] |
| HYPERION | Hospital mortality | BART | 0.19 | 0.77 | [0.68, 0.85] |
| HYPERION | 90-day CPC 1–2 | BART | 0.75 | 0.85 | [0.75, 0.93] |
| HYPERION | Hospital mortality | CausalForestDML | 0.19 | NA | NA |
| HYPERION | 90-day CPC 1–2 | CausalForestDML | 0.20 | NA | NA |
| PMAP | Hospital mortality | Neural network | No effect | 0.77 | [0.72, 0.82] |
| PMAP | Last mGCS = 6 | Neural network | No effect | 0.75 | [0.68, 0.81] |
| PMAP | Hospital mortality | XGBoost | 0.86 | 0.75 | [0.70, 0.80] |
| PMAP | Last mGCS = 6 | XGBoost | 0.70 | 0.76 | [0.70, 0.82] |
| PMAP | Hospital mortality | BART | 0.73 | 0.77 | [0.72, 0.82] |
| PMAP | Last mGCS = 6 | BART | 0.85 | 0.74 | [0.68, 0.79] |
| PMAP | Hospital mortality | CausalForestDML | 0.16 | NA | NA |
| PMAP | Last mGCS = 6 | CausalForestDML | 0.15 | NA | NA |
| MIMIC-IV | Hospital mortality | Neural network | 0.33 | 0.72 | [0.64, 0.80] |
| MIMIC-IV | Last mGCS = 6 | Neural network | No effect | 0.73 | [0.64, 0.81] |
| MIMIC-IV | Hospital mortality | XGBoost | 0.09 | 0.77 | [0.69, 0.84] |
| MIMIC-IV | Last mGCS = 6 | XGBoost | 0.89 | 0.74 | [0.65, 0.82] |
| MIMIC-IV | Hospital mortality | BART | 0.17 | 0.72 | [0.64, 0.79] |
| MIMIC-IV | Last mGCS = 6 | BART | 0.85 | 0.74 | [0.66, 0.82] |
| MIMIC-IV | Hospital mortality | CausalForestDML | 0.41 | NA | NA |
| MIMIC-IV | Last mGCS = 6 | CausalForestDML | 0.63 | NA | NA |

### Evidence of Heterogeneous Treatment Effects

No model demonstrated a significant CATE–TTM interaction by likelihood-ratio testing in any dataset or for any outcome (all p > 0.05; Table 2). The 95% confidence intervals for individual CATE estimates from CausalForestDML uniformly crossed zero across all datasets for the primary outcome of hospital mortality (Figure 2) and the secondary neurological outcome (eFigure 5), indicating no patient-level treatment effect that was statistically distinguishable from null.

**Figure 2.**
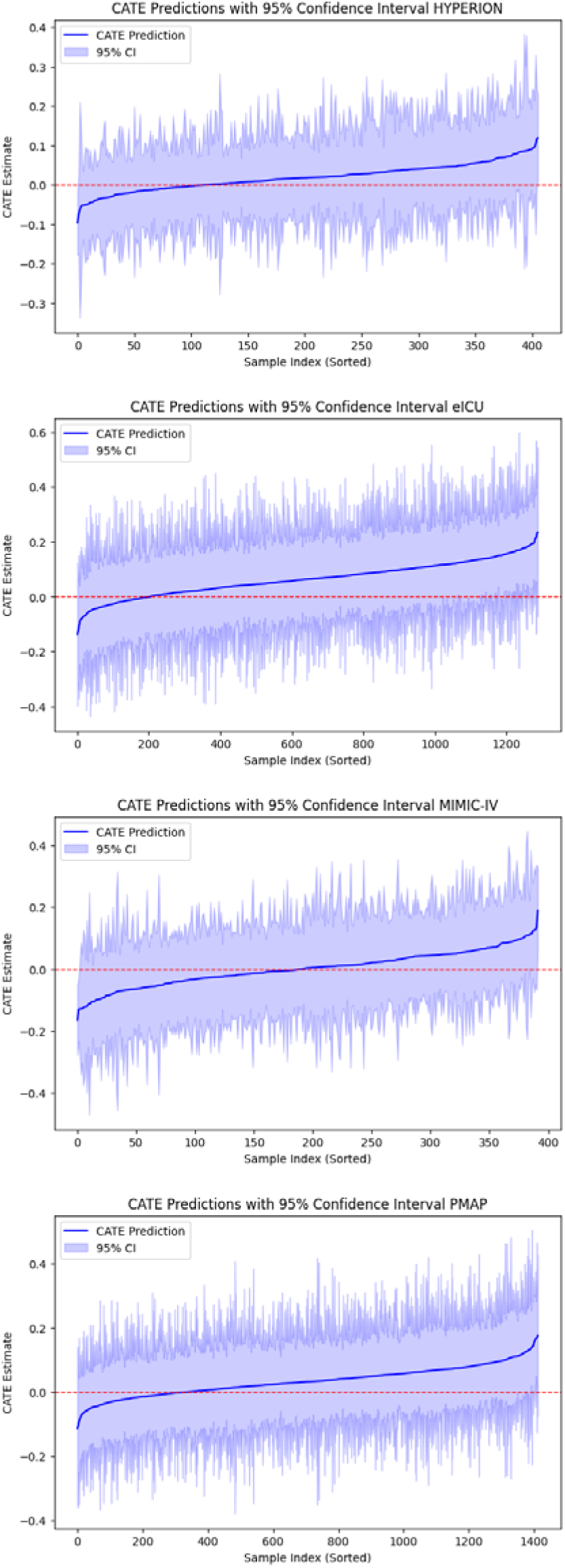
Conditional average treatment effect (CATE) predictions with 95% confidence intervals from CausalForestDML for hospital mortality in (A) HYPERION, (B) eICU-CRD, (C) PMAP, and (D) MIMIC-IV. All confidence intervals cross zero, indicating no patient-level treatment effect statistically distinguishable from null.

GATES analysis did not show significant treatment-effect heterogeneity in any dataset (all F-test p > 0.09; Figure 3, eTable 15). Quintile-specific treatment effects did not show a consistent monotonic pattern across datasets. In HYPERION, the pattern was inverse (ρ = −0.90, p = 0.037), further arguing against detectable HTE.

**Figure 3.**
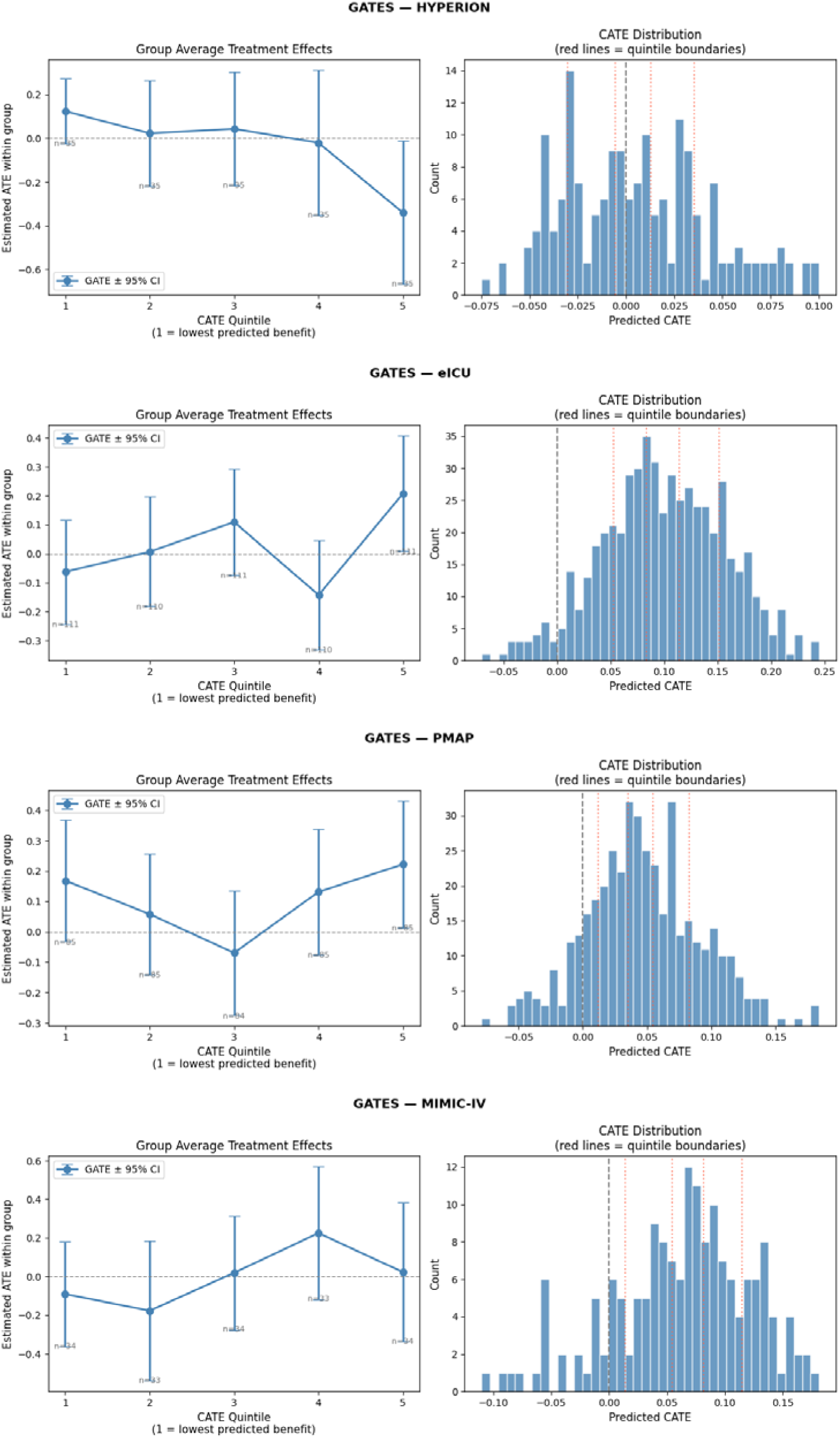
Group Average Treatment Effects (GATES) for hospital mortality. GATES are estimated within quintiles of predicted CATE from CausalForestDML across HYPERION, eICU-CRD, PMAP, and MIMIC-IV. Points are inverse-probability-weighted average treatment-effect estimates (unweighted ordinary least squares for HYPERION); error bars are 95% confidence intervals. The dashed line indicates the null. F-test p-value and Spearman ρ for monotonicity are annotated in each panel. Numeric results are provided in eTable 15.

SHAP analysis showed that TTM ranked among the least important features for outcome prediction across all datasets. In neural network models for the primary outcome of hospital mortality, TTM ranked 29th of 46 features (eICU-CRD), 51st of 222 (HYPERION), 93rd of 294 (MIMIC-IV), and 104th of 310 (PMAP) (eTable 6). Similar results were seen for the neurological outcome (eTable 5).

### Model Calibration

Calibration was assessed for both outcomes across all four datasets; eFigures 3–4 and eTables 3–4 show the full calibration results. Overall, calibration was adequate for the models used in CATE estimation, supporting the validity of the predicted probabilities from which treatment-effect estimates were derived.

## DISCUSSION

Applying causal machine learning across one RCT and three observational datasets encompassing 4,507 cardiac-arrest patients, we found no evidence of heterogeneous treatment effects for TTM. This finding was consistent across multiple metalearner architectures, base models, datasets, and outcome definitions.

### Interpretation in Context

A prevailing hypothesis has been that discrepant TTM trial results might be explained by differential benefit in patient subgroups that conventional pre-specified analyses fail to detect.^7,19^ Our causal machine-learning approach, which can model complex non-linear interactions without pre-specifying subgroups, does not support this hypothesis. This aligns with the individual patient data meta-analysis of TTM1 and TTM2, which found no interaction between hypothermia and pre-specified subgroup variables including age, sex, initial rhythm, time to ROSC, and circulatory shock.^5^

The causal conclusions of this study rest primarily on the GATES analysis, with the CATE interaction test and CausalForestDML confidence intervals providing additional support. SHAP findings were used as descriptive evidence only, as SHAP reflects predictive importance rather than causal effect on outcomes.^21^ Overall, we interpret the combined evidence of the four evaluation approaches as indicating a lack of HTE for TTM.

### Strengths

This study has several strengths. The multi-dataset design, incorporating both randomized and observational data in heterogeneous clinical settings and geographies (France, two U.S. academic centers, and a multicenter U.S. registry), strengthens generalizability. The use of multiple metalearner architectures and base models—three S-learners and one forest-based R-learner— reduces dependence on any single analytical approach. The convergence of four complementary HTE assessment methods (LR interaction test, GATES, CausalForestDML confidence intervals, and SHAP analysis) adds robustness to the negative finding. Feature selection guided by the HYPERION RCT provides clinical grounding and reduces the risk that feature-selection artifacts drive the null result.

### Limitations

Several limitations should be discussed. First, the validity of causal machine-learning estimates depends on conditional ignorability, which is satisfied by design in HYPERION but untestable in the three observational cohorts. Several clinically important factors were not captured in our covariate sets, including time-to-target temperature,^20^ withdrawal-of-life-sustaining-treatment practices, site-level TTM protocols, and the treating clinician’s prognostic assessment at TTM initiation. CATE estimates in the observational cohorts should therefore be interpreted as conditional associations rather than unbiased causal effects. CausalForestDML partially addresses this through propensity-based orthogonalization, but this correction is only as good as the measured covariates. However, the null HTE finding was consistent between HYPERION (where confounding is absent by design) and the observational datasets, arguing against residual confounding as a primary driver of the negative result.

Second, TTM was defined differently across datasets. PMAP used a temperature-based proxy (sustained hypothermia <36°C for >12 hours), which cannot separate therapeutic cooling from spontaneous post-arrest hypothermia. Temperature trajectories support the fidelity of TTM labels in HYPERION, eICU-CRD, and MIMIC-IV, and indicate a reasonable though less precise proxy in PMAP (Figure 4). Any residual misclassification would bias CATE estimates toward the null, but the null result held across all four datasets, including the three with high-fidelity labels, arguing against misclassification as the driver.

**Figure 4.**
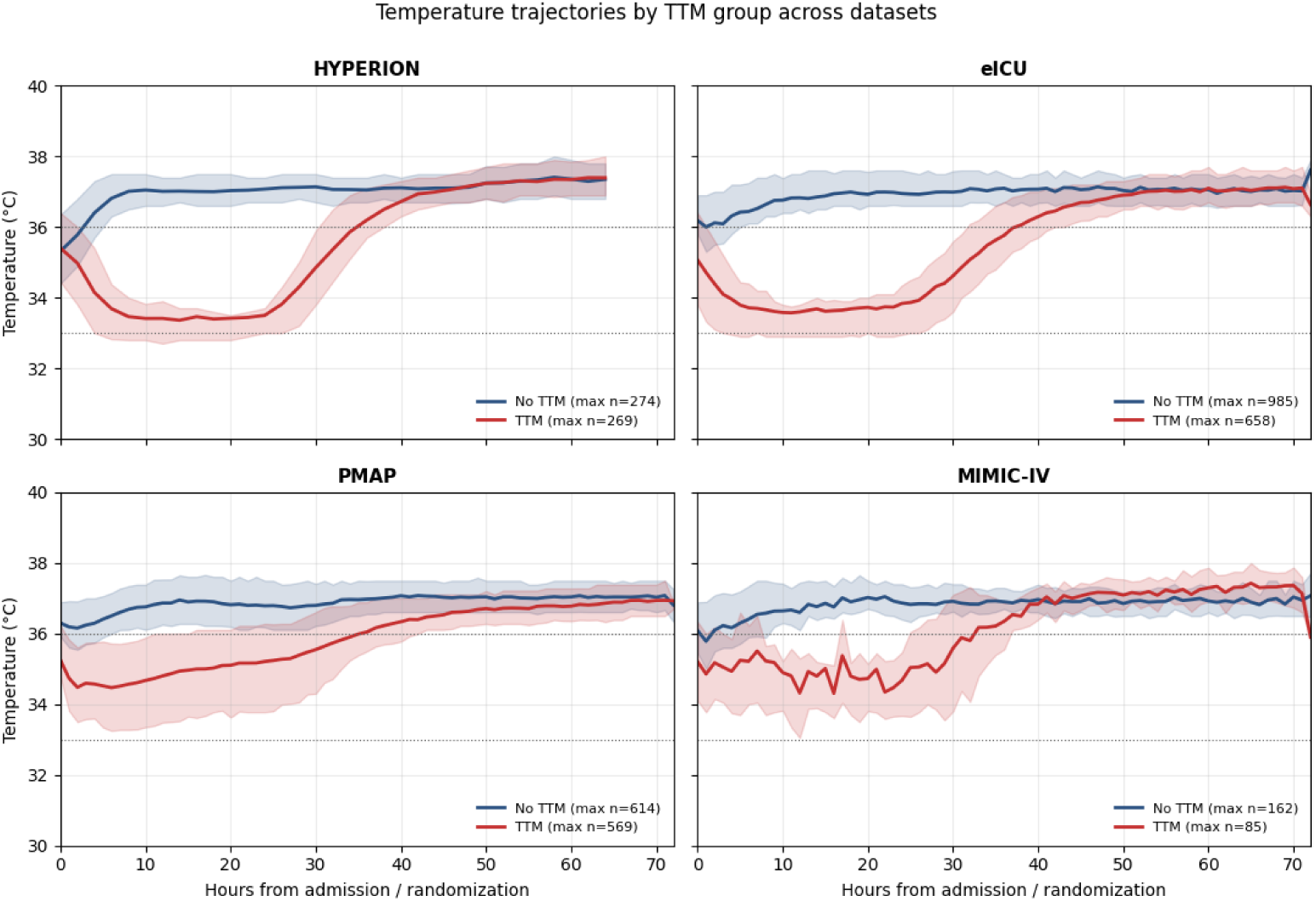
Temperature trajectories by TTM group across datasets over the first 72 hours from randomization (HYPERION) or admission (eICU-CRD, PMAP, MIMIC-IV). Solid lines show group mean temperature; shaded bands show the interquartile range. Dashed horizontal lines indicate the 33°C and 36°C clinical targets used in the major TTM trials. Sample sizes refer to the maximum number of patients contributing a temperature measurement in any hourly bin. TTM = targeted temperature management.

Third, the primary outcome was death at discharge across all datasets, improving comparability for the main analysis. Our secondary neurological outcome was less harmonized: HYPERION used 90-day CPC, while the observational cohorts used the last recorded motor GCS at discharge. Because motor GCS is not a validated long-term functional outcome measure, this secondary endpoint is less directly comparable across cohorts. In addition, assigning deceased patients a motor GCS of 1 in the observational datasets partially combines mortality with neurological outcome. These differences may have added noise to the secondary analyses but are unlikely to explain the overall null HTE findings.

Fourth, detecting heterogeneous treatment effects requires substantially larger sample sizes than detecting average treatment effects, because the signal of interest—differential response across patient subgroups—is inherently weaker than the overall treatment effect. To assess this, we conducted post-hoc power simulations with 10%, 20%, and 30% treatment-effect benefits in subgroups comprising 20%, 30%, or 50% of patients. Power was high for larger treatment-effect sizes (≥0.2 absolute risk difference), exceeding 0.90 across most datasets, suggesting large HTE is unlikely to have been missed. However, power was limited for smaller effect sizes (10 percentage-point benefit) in smaller subgroups (20% of patients), where power ranged from 0.16 to 0.50. Full simulation results are provided in eTables 7–14.

Last, missing data were handled using k-nearest-neighbor imputation across all datasets, without formal assessment of the missingness mechanism or sensitivity analyses using alternative strategies (e.g., multiple imputation, complete-case analysis). The robustness of our findings to alternative missing-data approaches is therefore unverified, although differential missingness between TTM and non-TTM groups was limited (Results; eTables 16–19), arguing against missing-not-at-random as a major source of bias.

### Clinical Implications

If the null HTE finding is confirmed in larger pooled analyses, it would weaken the case for personalized TTM strategies. Instead, these results support redirecting efforts toward other neuroprotective strategies after cardiac arrest and toward optimizing other modifiable elements of post-arrest care, including hemodynamic management, seizure prevention, and structured neuroprognostication protocols. It remains possible, however, that HTE for TTM operates through mechanisms not captured in early clinical features—for example, differential responses mediated by biomarker profiles, genetic susceptibility, or the timing and speed of cooling initiation—and that future studies incorporating these dimensions may yet reveal differential benefit.

## CONCLUSIONS

Using causal machine learning in four diverse datasets, we found no evidence that heterogeneous treatment effects explain the inconclusive results of TTM trials in cardiac arrest. These findings, which build on recent individual patient data meta-analyses, should be interpreted with caution given the limitations of sample size, unmeasured confounding in observational data, and treatment-ascertainment heterogeneity. Nonetheless, the convergence of null findings across randomized and observational data, multiple analytical approaches, and diverse patient populations provides meaningful evidence against the hypothesis that identifiable patient subgroups derive differential benefit from TTM as currently applied.

## Data Availability

All data produced in the present work are in the manuscript

## Author Contributions

MBR: study design, data analysis, manuscript drafting. IKL: data extraction, analysis support. CH: data extraction, analysis support. RP: analysis support, critical revision. JBL: HYPERION data provision, critical revision. RDS: study conception, study design, supervision, data analysis, critical revision, final approval. All authors read and approved the final manuscript.

## Copyright Form Disclosures

Each author has completed and submitted the Copyright Transfer Agreement.

