## Supplementary Appendix for "Heterogeneous Treatment Effect for Targeted Temperature Management After Cardiac Arrest: A Causal Machine Learning Analysis"

| **Section** | **Content** |
| --- | --- |
| S.1 | Data Processing Pipeline (eFigure 1) |
| S.2 | Propensity Score Diagnostics (eFigure 2) |
| S.3 | Model Calibration Plots (eFigures 3-4) |
| S.4 | CATE 95% Confidence Intervals for Neurologic Outcome (eFigure 5) |
| S.5 | Patient Selection Flowchart (eFigure 6) |
| S.6 | Patient and Dataset Selection (eTable 1) |
| S.7 | Causal Machine Learning Overview — Meta-Learners (eTable 2) |
| S.8 | Model Calibration Statistics (eTables 3-4) |
| S.9 | Feature Importance — SHAP Values (eTables 5-6) |
| S.10 | Power and Sensitivity Analyses (eTables 7-14) |
| S.11 | GATES Numeric Results (eTable 15) |
| S.12 | Feature List and Per-Dataset Missing Data Report (eTables 16-19) |
| S.13 | STROBE Checklist (eTable 20) |

### S.1 Data Processing

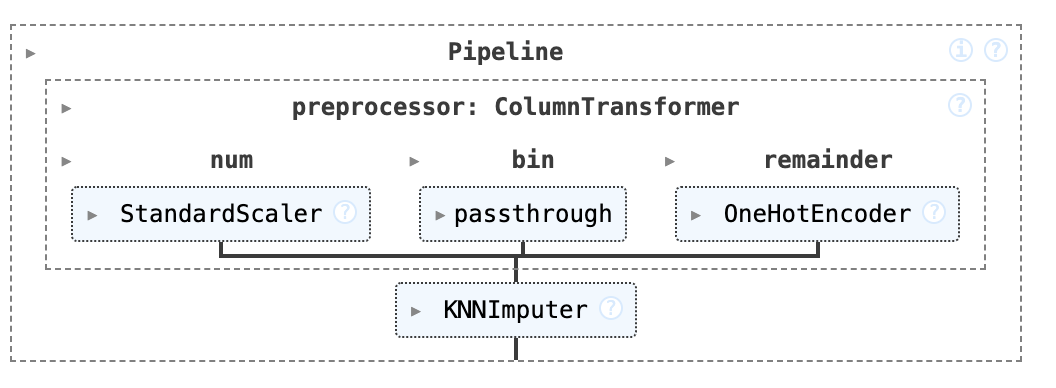

**eFigure 1** – Data processing pipeline. Numerical variables were standard-scaled, binary variables were passed through unchanged, and categorical variables were one-hot encoded; the resulting feature vector was then imputed using a KNN imputer for missing values.

### S.2 Propensity Score Diagnostics

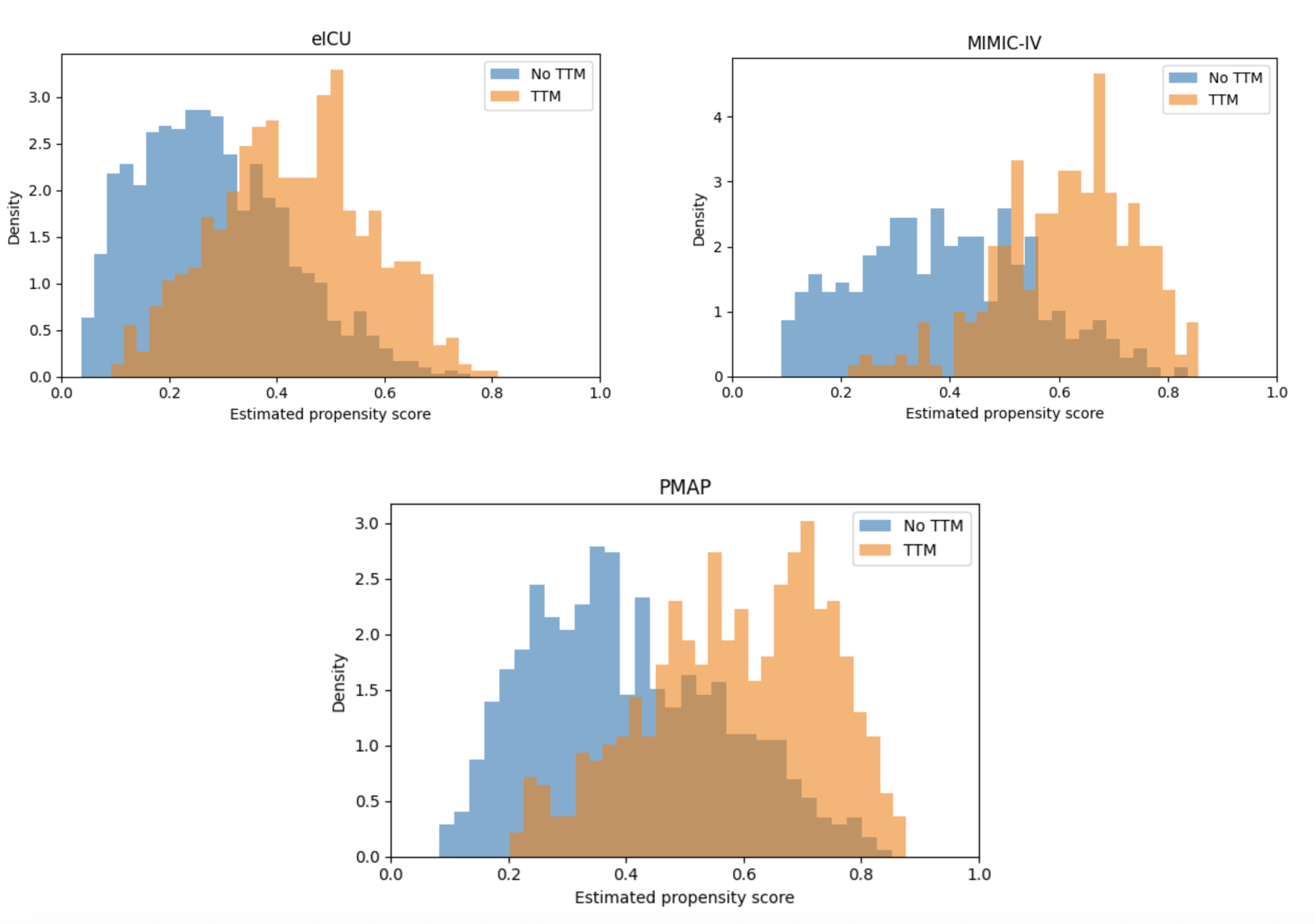

**eFigure 2** – Overlap density plots for propensity scores in the observational datasets. The x-axis shows the propensity score and the y-axis shows probability density. Patients with higher propensity scores were more likely to receive TTM, as expected; substantial overlap between groups indicates adequate positivity. No major positivity violations were detected (the proportion of patients with propensity score < 0.05 or > 0.95 was less than 0.3% in all datasets).

### S.3 Model Calibration Plots

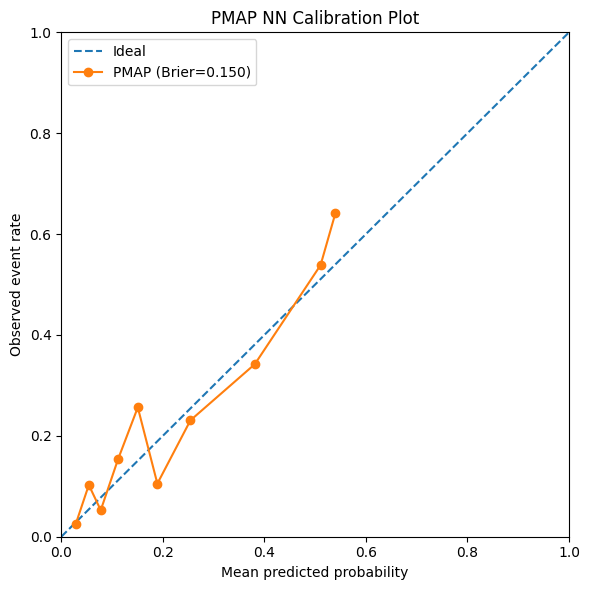

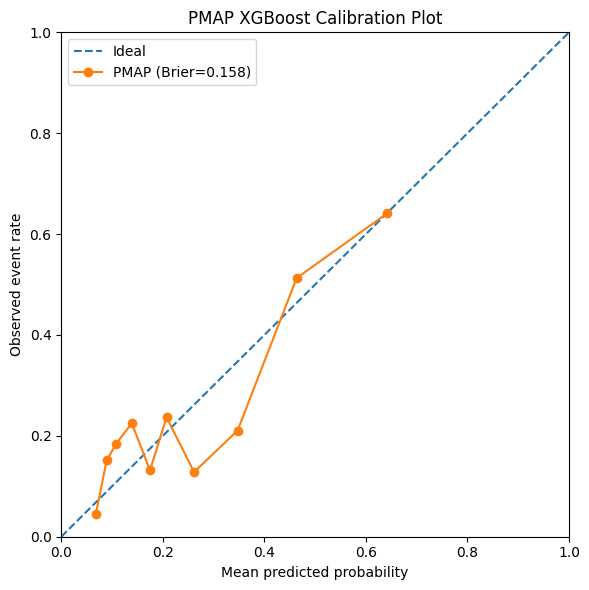

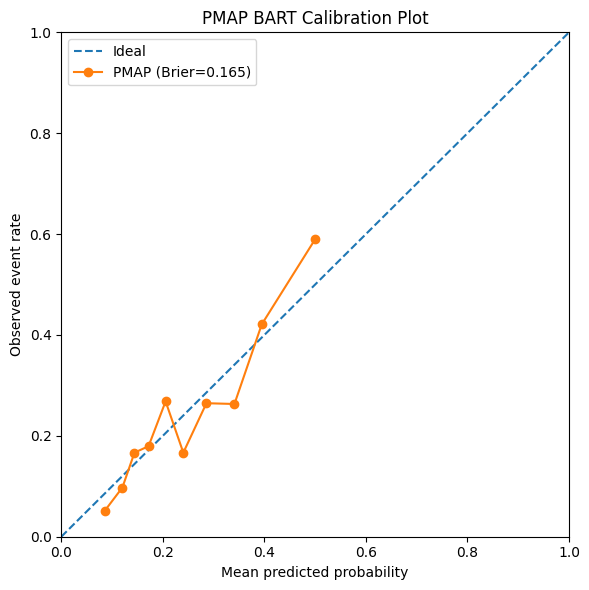

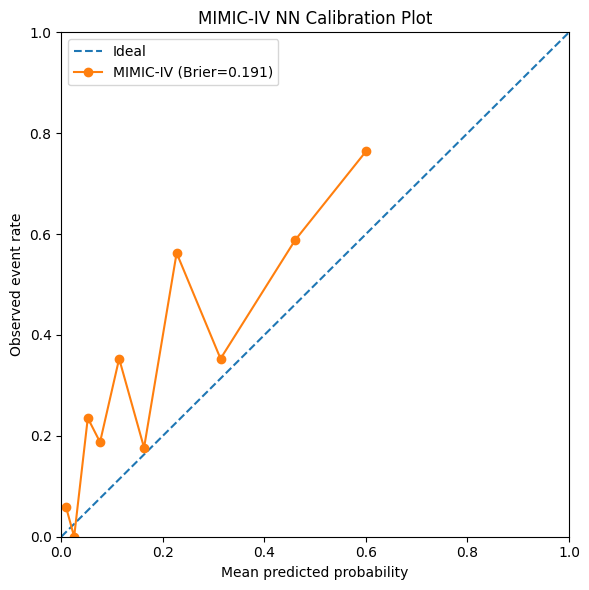

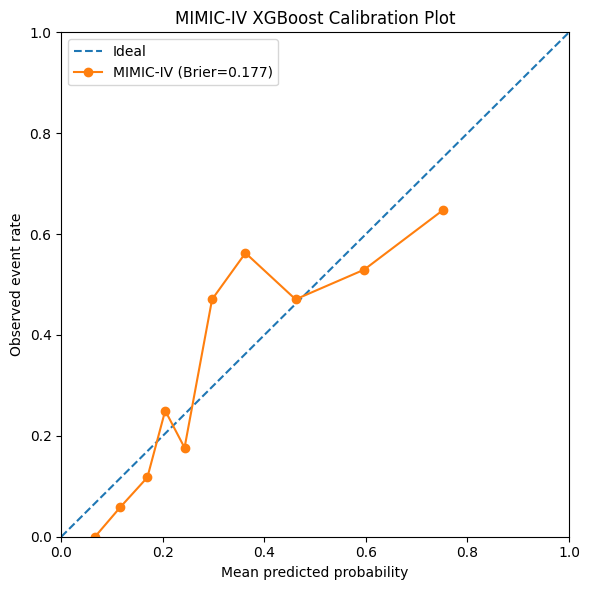

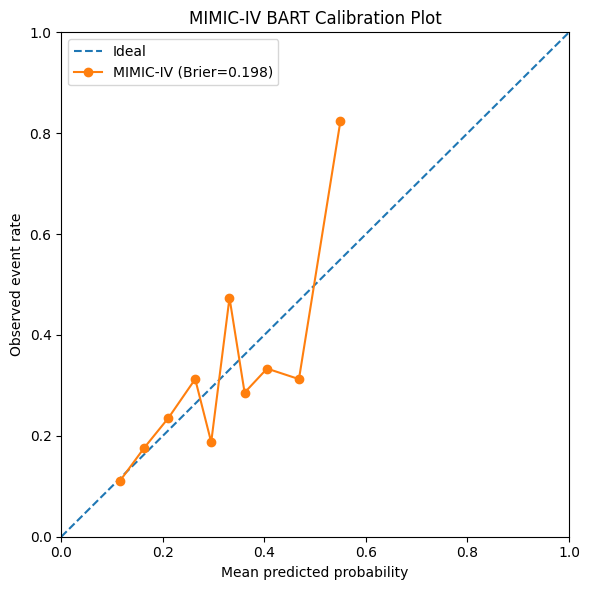

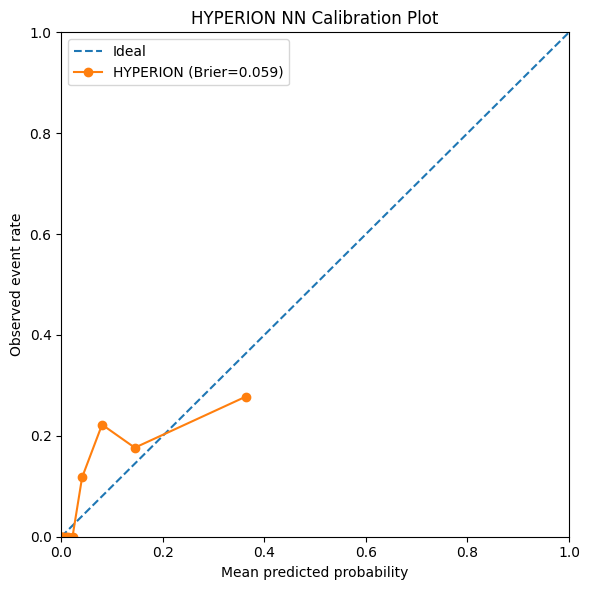

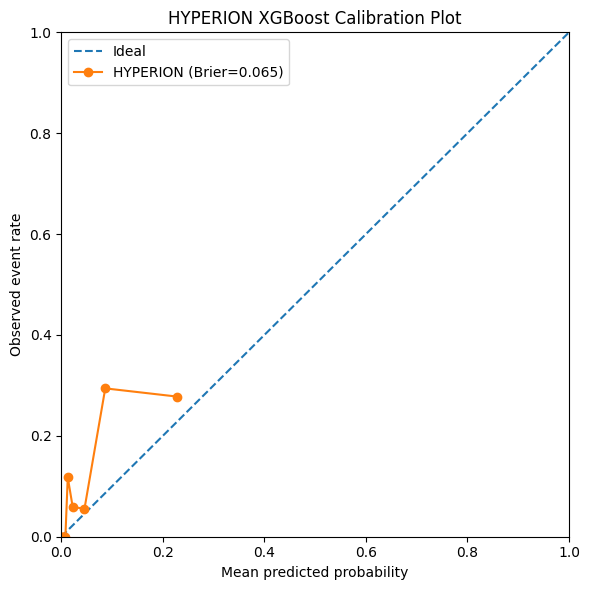

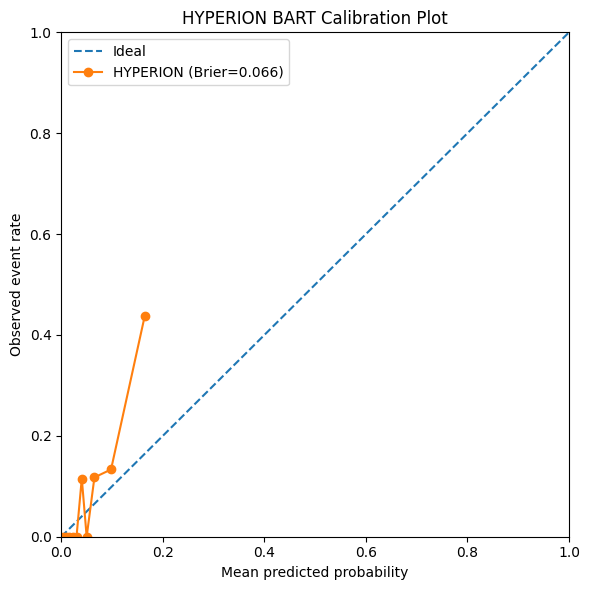

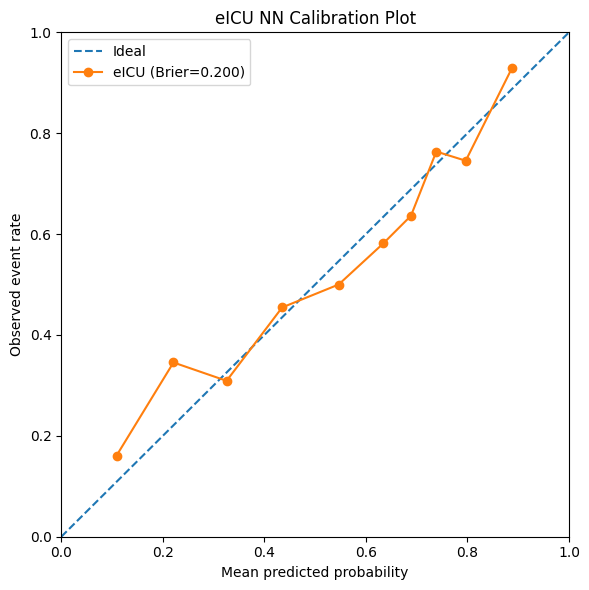

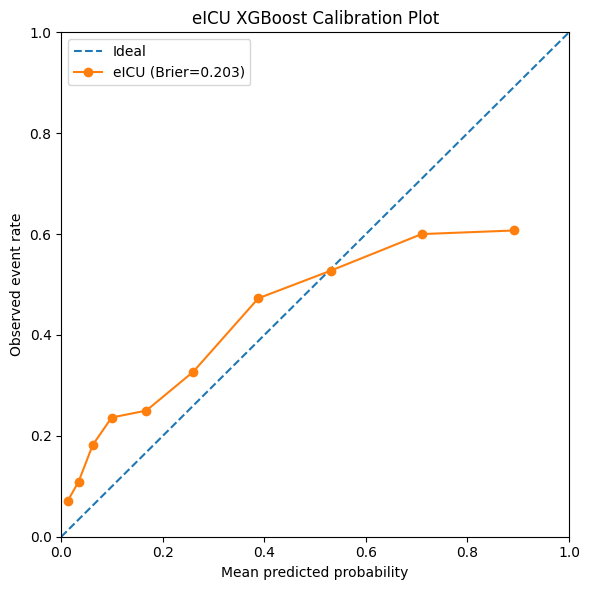

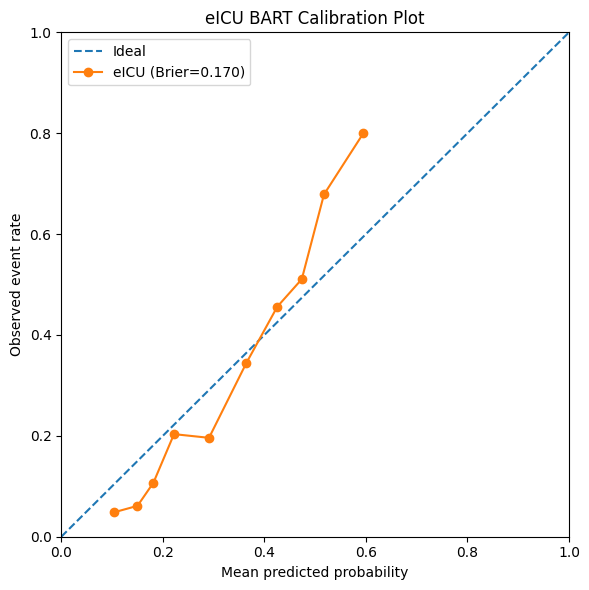
**eFigure 3** – Calibration plots for S-learner models predicting the neurological outcome. Calibration was better in datasets with larger sample sizes.

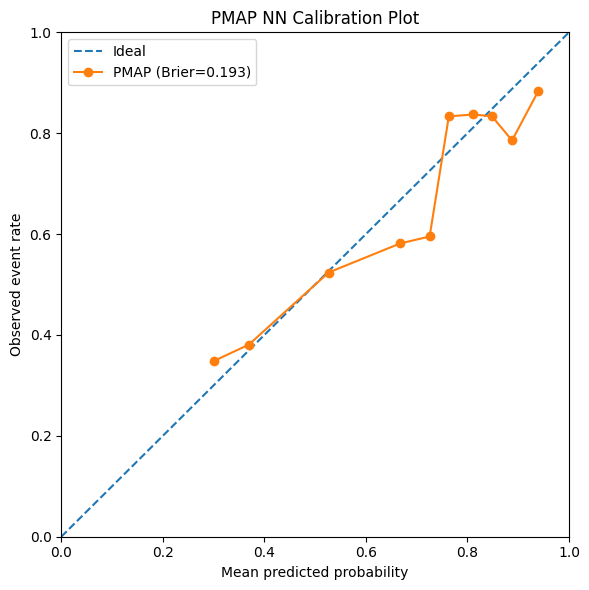

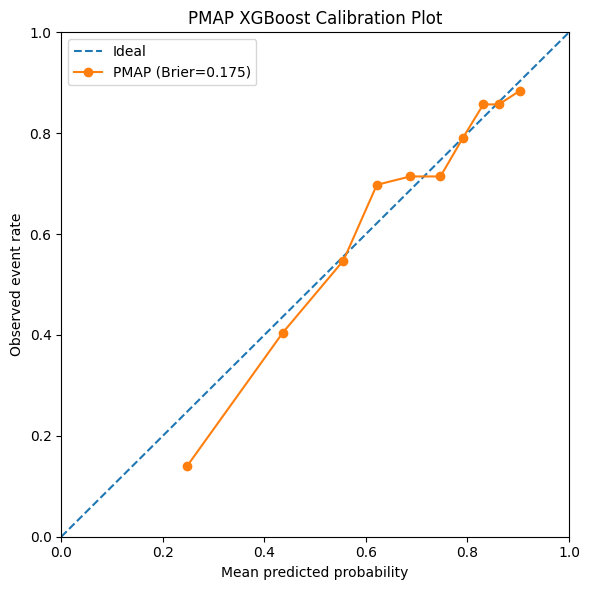

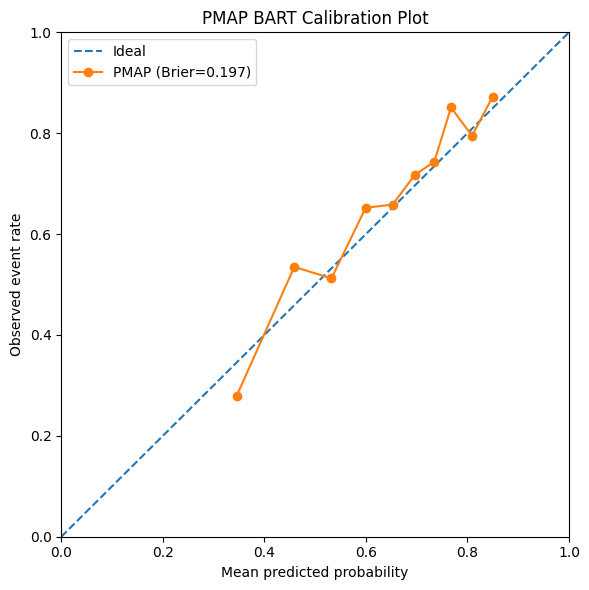

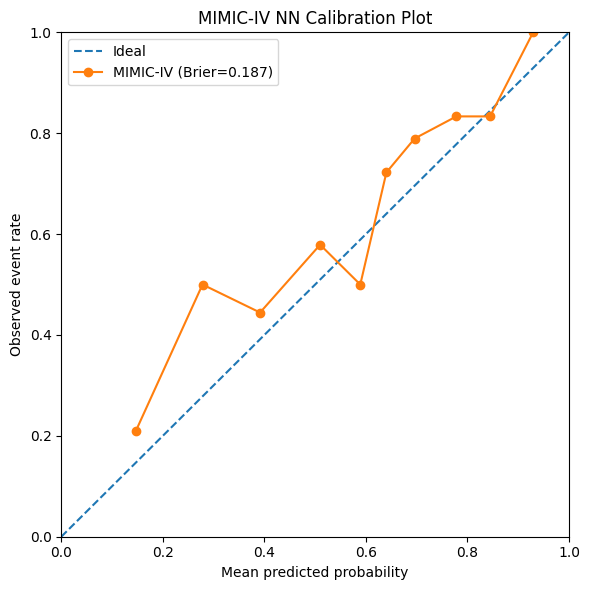

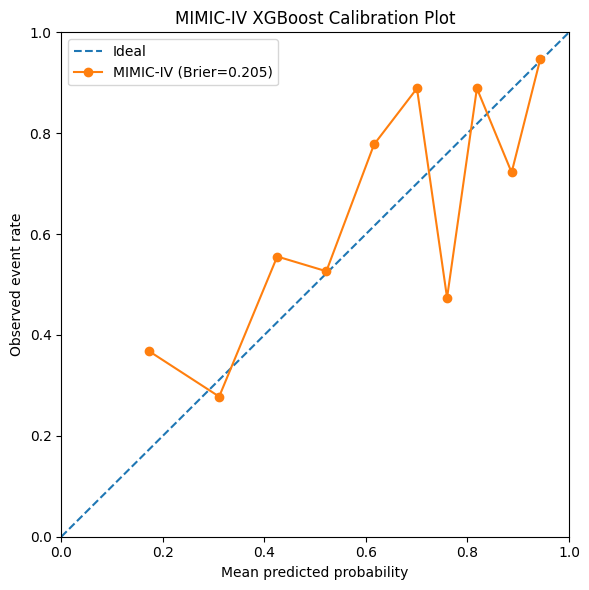

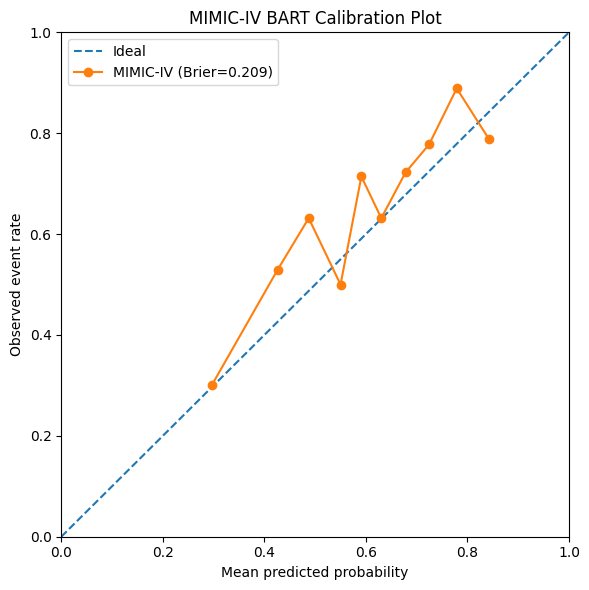

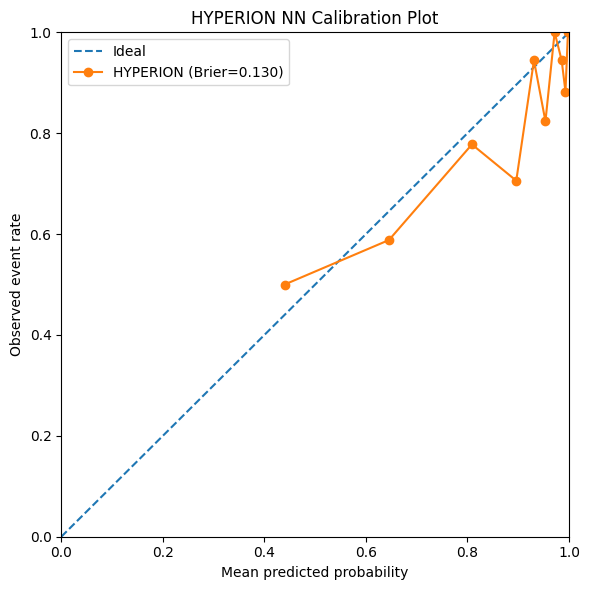

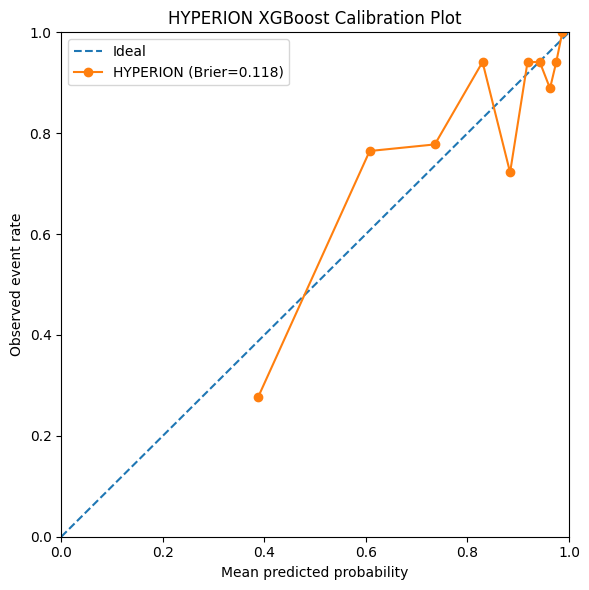

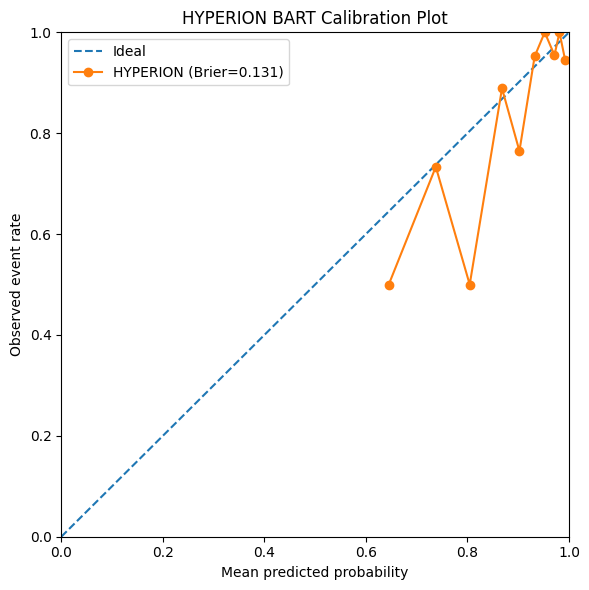

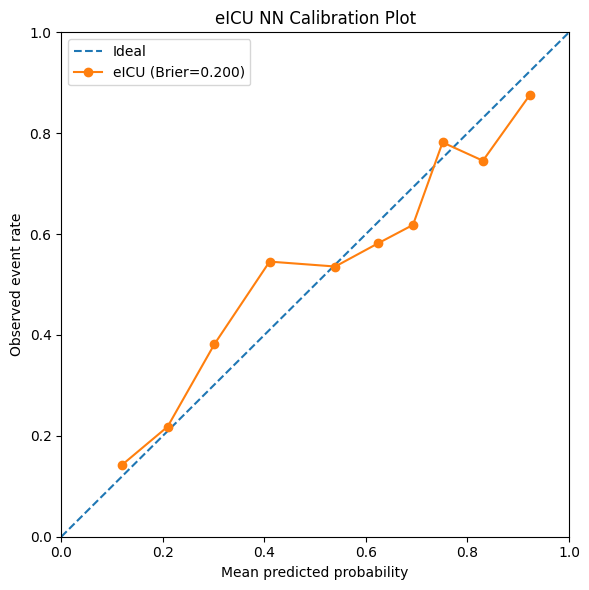

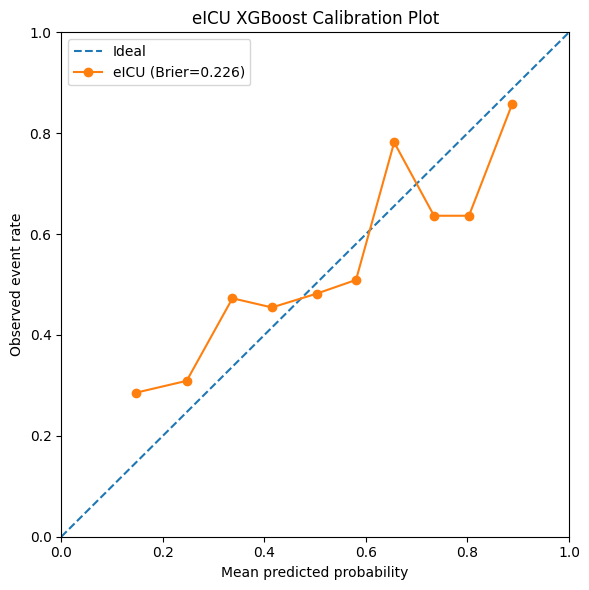

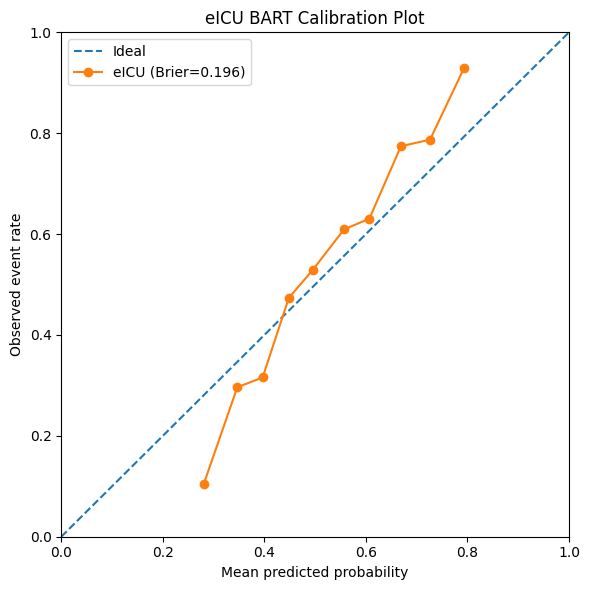

**eFigure 4** – Calibration plots for S-learner models predicting hospital mortality. Calibration was better in datasets with larger sample sizes.

### S.4 CATE 95% Confidence Intervals for Neurologic Outcomes

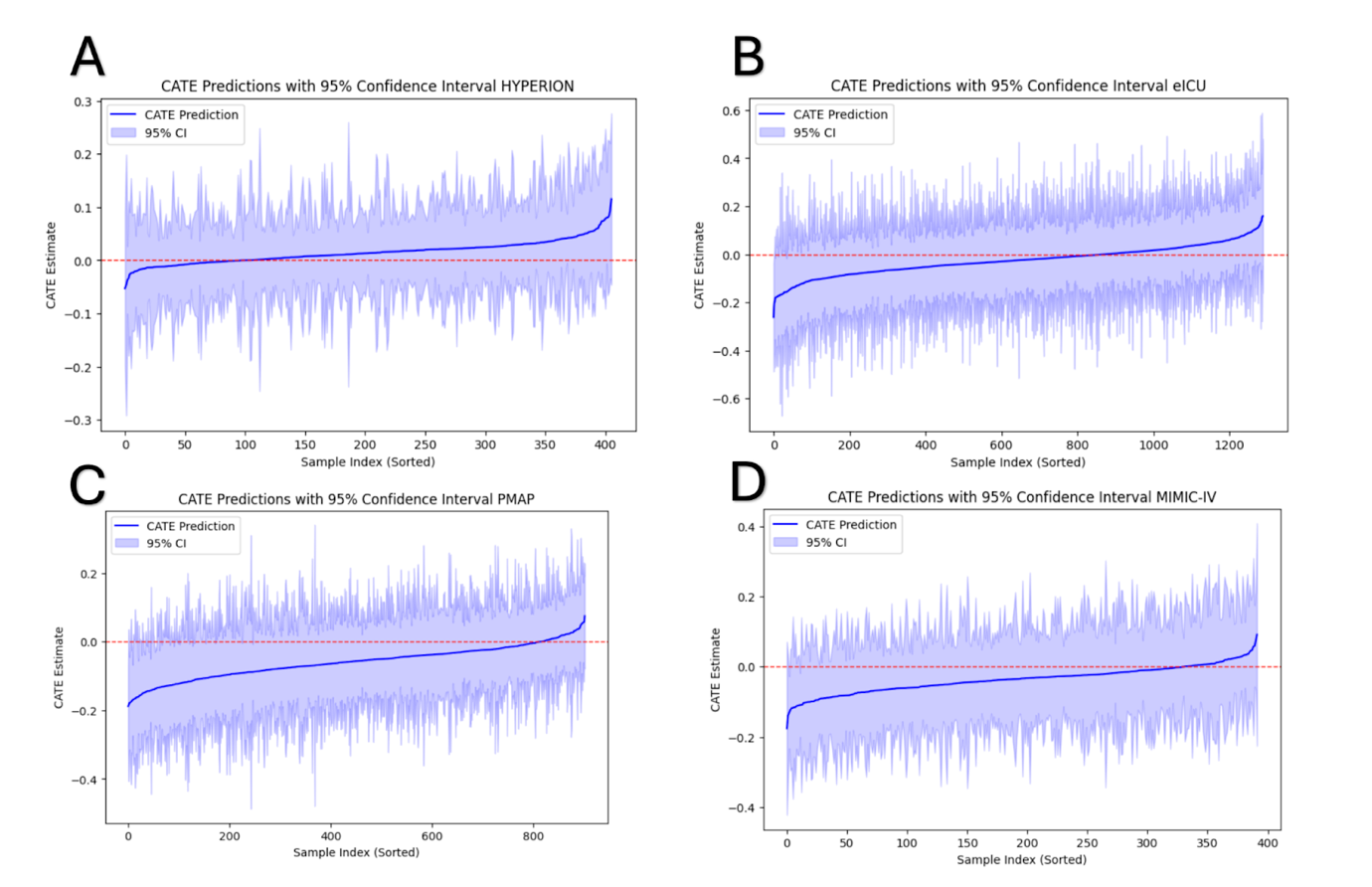

**eFigure 5 –** CATE predictions with 95% confidence intervals from CausalForestDML models for neurologic outcomes in (A) HYPERION, (B) eICU-CRD, (C) PMAP, and (D) MIMIC-IV.

**S.5 Patient Selection Flowchart**

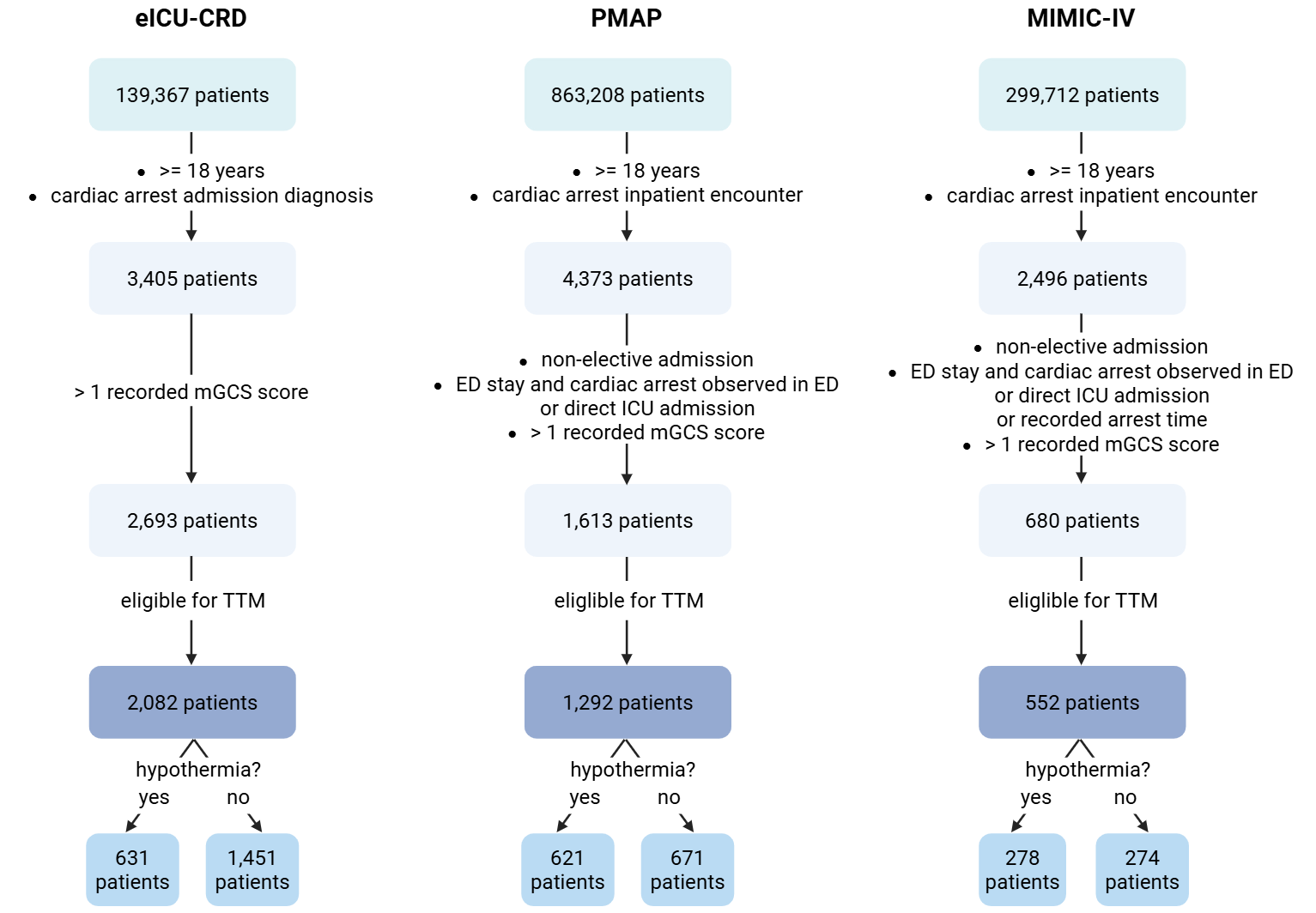

**eFigure 6** – Patient selection flowchart for the observational cohorts. Eligibility required a first measured motor GCS < 6 and an mGCS measurement available within 6 hours of cardiac arrest**.**

### S.6 Patient and Dataset Selection

| **Dataset** | **Study type** | **Geography / sites** | **Data years** | **Analytic sample (TTM / no TTM)** | **Inclusion criteria** | **Primary outcome(s)** | **Data access** |
| --- | --- | --- | --- | --- | --- | --- | --- |
| **HYPERION** | Randomized controlled trial | France; 26 ICUs | 2014–2018 | 581 total (284 TTM / 297 normothermia) | Adults ≥18 yrs with OHCA or IHCA due to non-shockable rhythm; GCS <8 at ICU admission; randomized to TTM 33°C vs. targeted normothermia 37°C | In-hospital mortality (primary); 90-day CPC score (favorable = CPC 1–2), (secondary) | Published trial; ClinicalTrials.gov NCT02057835 |
| **eICU-CRD** v2.0 | Multicenter retrospective cohort | United States; ~200 ICUs | 2014–2015 | 2,082 total (631 TTM / 1,451 no TTM) | Adults ≥18 yrs with cardiac arrest admission; first recorded mGCS < 6 within 6 hours of arrest; mGCS measurement available | In-hospital mortality (primary); last recorded mGCS (favorable = mGCS 6) (secondary) | PhysioNet credentialed access; IRB waiver (de-identified) |
| **PMAP** (Johns Hopkins) | Single-center retrospective cohort | United States; Johns Hopkins Hospital, Baltimore | 2016–2024 | 1,292 total (621 TTM / 671 no TTM) | Adults ≥18 yrs with cardiac arrest admission to Johns Hopkins ICU; first recorded mGCS < 6 within 6 hours of arrest; mGCS measurement available | In-hospital mortality (primary); last recorded mGCS (favorable = mGCS 6) (secondary) | JHU IRB approval; institutional data use agreement |
| **MIMIC-IV** v2.2 | Single-center retrospective cohort | United States; Beth Israel Deaconess Medical Center, Boston | 2008–2022 | 552 total (278 TTM / 274 no TTM) | Adults ≥18 yrs with cardiac arrest ICD code; first recorded mGCS < 6 within 6 hours of arrest; mGCS measurement available | In-hospital mortality (primary); last recorded mGCS (favorable = mGCS 6) (secondary) | PhysioNet credentialed access; IRB waiver (de-identified) |

**eTable 1** – Dataset description and patient selection criteria.

### S.7 Causal Machine Learning Overview

Several causal machine learning approaches have been developed to identify HTE, most prominently the family of meta-learners: S-, T-, X-, and R-learners [8], [9] (see eTable 2). The **S-learner** (single-learner) fits one outcome model that includes treatment as a covariate, then estimates the conditional average treatment effect (CATE) as the difference between predictions with treatment set to 1 versus 0. Flexible base learners such as Bayesian Additive Regression Trees (BART) are commonly used in this framework [15]. The **T-learner** (two-learner) fits two separate outcome models, one on treated and one on control units, and estimates the CATE as the difference between their predicted counterfactual outcomes. The **X-learner** extends the T-learner by imputing individual-level treatment effects for each arm using the opposite arm's model, fitting a second-stage model to these pseudo-outcomes, and combining the two CATE estimates with propensity-score weights; this makes it particularly efficient when treatment groups are highly imbalanced, as is often the case for targeted temperature management (TTM), where patients with higher GCS are less likely to receive the intervention. The **R-learner** [12] residualizes both the outcome and the treatment using cross-fitted nuisance models (following Robinson's decomposition) and then minimizes the R-loss to recover the CATE function, a construction that gives it Neyman orthogonality with respect to nuisance estimation errors [9].

CausalForestDML, the primary causal estimator used in this study, belongs to the same double machine learning (DML) family as the R-learner and can be viewed as a forest-based R-learner: it cross-fits outcome and propensity nuisance models and solves a local version of the R-loss within the leaves of a generalized random forest, yielding pointwise asymptotically normal CATE estimates with valid confidence intervals. It should be distinguished from genuinely doubly robust estimators such as EconML's ForestDRLearner or AIPW-based DR-learners, which use doubly robust (augmented inverse-propensity-weighted) moment conditions and remain consistent if *either* the outcome or the propensity model is correctly specified. We selected CausalForestDML for its combination of nonparametric flexibility, honest inference, and strong empirical performance on observational ICU data, while acknowledging that a DR-learner variant would offer complementary robustness properties and could be considered in sensitivity analyses.

| **Meta-Learner** | **Estimator** | **CATE** |
| --- | --- | --- |
| S-learner | u(x)= E(Y\|X, T) | CATE(x) = u(x\|T=1)-u(X\|t=0) |
| T-learner | u1(x)= E(Y\|X, T = 1)  u0(x)= E(Y\|X, T = 0) | CATE(x) = u1(x)-u0(X) |
| X-learner | D0(x)= Y-u0(x)  D1(x)= Y-u1(x)  t1(x)= E(D1\|X)  t0(x)= E(D0\|X) | CATE(x) = e(x)t0(x)- (1-e(x))t1(x) |
| R-learner | Yi- u(x) = (Ti-e(x))t(x)+ei | CATE(x) = mint{Yi-u(x)-(Ti-e(x))t(x)} |
| DR-learner (CausalForestDML) | ê(x) = cross-fitted E(Y\|X), ê(x) = cross-fitted P(T=1\|X) | CATE(x) = argmin_τ Σ[(Ỹᵢ - τ(Xᵢ)T̃ᵢ)²] where Ỹ = Y - ê(x), T̃ = T - e(x) |

**eTable 2** – Meta-learner frameworks for causal inference. e(x) = P(T=1|X) is the propensity score, T is the treatment variable, X are covariates, and Y is the outcome. CausalForestDML is a forest-based R-learner in the double machine learning (DML) family: outcome and propensity nuisance functions are cross-fitted, yielding Neyman-orthogonal CATE estimates whose first-order bias from nuisance misspecification vanishes. It should be distinguished from AIPW-based DR-learners (e.g., EconML’s ForestDRLearner), which are doubly robust in the strict sense of remaining consistent if either nuisance model is correctly specified.

### S.8 Model Calibration Statistics

| **dataset** | **outcome** | **model** | **n** | **event_rate** | **brier** | **calibration_intercept** | **calibration_slope** |
| --- | --- | --- | --- | --- | --- | --- | --- |
| PMAP | Neurologic | NN | 387 | 0.245478 | 0.149878 | 0.098179 | 0.992777 |
| PMAP | Neurologic | XGBoost | 387 | 0.245478 | 0.157853 | -0.134294 | 0.888028 |
| PMAP | Neurologic | BART | 387 | 0.245478 | 0.164632 | 0.140442 | 1.155819 |
| MIMIC-IV | Neurologic | NN | 168 | 0.327381 | 0.19063 | 0.580602 | 0.797482 |
| MIMIC-IV | Neurologic | XGBoost | 168 | 0.327381 | 0.17693 | 0.199684 | 1.088321 |
| MIMIC-IV | Neurologic | BART | 168 | 0.327381 | 0.198275 | 0.146132 | 1.117615 |
| HYPERION | Neurologic | NN | 175 | 0.08 | 0.059075 | 0.013774 | 0.891417 |
| HYPERION | Neurologic | XGBoost | 175 | 0.08 | 0.059432 | 0.133516 | 0.839987 |
| HYPERION | Neurologic | BART | 175 | 0.08 | 0.065559 | 2.72419 | 1.855952 |
| eICU | Neurologic | NN | 553 | 0.338156 | 0.173716 | -0.054466 | 0.760966 |
| eICU | Neurologic | XGBoost | 553 | 0.338156 | 0.202565 | -0.186212 | 0.485625 |
| eICU | Neurologic | BART | 553 | 0.338156 | 0.177563 | 0.339948 | 1.397203 |

**eTable 3** – Calibration Table containing brier scores, slopes and intercepts for neurologic outcomes in datasets.

| **dataset** | **outcome** | **model** | **n** | **event_rate** | **brier** | **calibration_intercept** | **calibration_slope** |
| --- | --- | --- | --- | --- | --- | --- | --- |
| PMAP | Mortality | NN | 424 | 0.660377 | 0.193012 | 0.024036 | 0.771753 |
| PMAP | Mortality | XGBoost | 424 | 0.660377 | 0.174861 | -0.133174 | 1.145311 |
| PMAP | Mortality | BART | 424 | 0.660377 | 0.196955 | 0.025935 | 1.126102 |
| MIMIC-IV | Mortality | NN | 184 | 0.641304 | 0.186645 | 0.344551 | 0.94413 |
| MIMIC-IV | Mortality | XGBoost | 184 | 0.641304 | 0.205168 | 0.267104 | 0.639798 |
| MIMIC-IV | Mortality | BART | 184 | 0.641304 | 0.209006 | 0.22324 | 0.947963 |
| HYPERION | Mortality | NN | 175 | 0.817143 | 0.130064 | 0.130568 | 0.612496 |
| HYPERION | Mortality | XGBoost | 175 | 0.817143 | 0.117571 | 0.14825 | 0.836016 |
| HYPERION | Mortality | BART | 175 | 0.817143 | 0.1307 | 0.13352 | 0.629031 |
| eICU | Mortality | NN | 553 | 0.542495 | 0.199685 | 0.037307 | 0.810618 |
| eICU | Mortality | XGBoost | 553 | 0.542495 | 0.225758 | 0.089208 | 0.639133 |
| eICU | Mortality | BART | 553 | 0.542495 | 0.19607 | 0.010032 | 1.74057 |

**eTable 4** – Calibration Table containing brier scores, slopes and intercepts for hospital mortality outcomes in datasets.

### S.9 Feature Importance

| **Dataset** | **TTM Shap** | **TTM Rank out of all features** |
| --- | --- | --- |
| eICU | .0170 | 29/46 |
| HYPERION | .0002 | 171/222 |
| MIMIC-IV | .0033 | 92/294 |
| PMAP | .0000 | 106/310 |

**eTable 5** – SHAP [21] values for TTM in each dataset (neural network models, neurological outcome), with the rank of TTM among all features. TTM was a low-importance feature for outcome prediction across all datasets.

| **Dataset** | **TTM Shap** | **TTM Rank out of all features** |
| --- | --- | --- |
| eICU | .0100 | 29/46 |
| HYPERION | .0009 | 51/222 |
| MIMIC-IV | .0008 | 93/294 |
| PMAP | .0000 | 104/310 |

**eTable 6** – SHAP values for TTM in each dataset (neural network models, primary outcome of hospital mortality), with the rank of TTM among all features. Results mirror those of eTable 5: TTM was a low-importance feature for outcome prediction across all datasets.

### S.10 Power and Sensitivity Analysis

| **Dataset** | **N** | **TTM prevalence** | **Responsive subgroup (%)** | **Baseline risk** | **Assumed absolute treatment effect** | **Simulations** | **Power** |
| --- | --- | --- | --- | --- | --- | --- | --- |
| HYPERION | 581 | 0.489 | 20 | 0.943 | 0.1 | 2,000 | 0.502 |
| HYPERION | 581 | 0.489 | 30 | 0.943 | 0.1 | 2,000 | 0.732 |
| HYPERION | 581 | 0.489 | 50 | 0.943 | 0.1 | 2,000 | 0.913 |
| HYPERION | 581 | 0.489 | 20 | 0.943 | 0.2 | 2,000 | 0.496 |
| HYPERION | 581 | 0.489 | 30 | 0.943 | 0.2 | 2,000 | 0.741 |
| HYPERION | 581 | 0.489 | 50 | 0.943 | 0.2 | 2,000 | 0.915 |
| HYPERION | 581 | 0.489 | 20 | 0.943 | 0.3 | 2,000 | 0.493 |
| HYPERION | 581 | 0.489 | 30 | 0.943 | 0.3 | 2,000 | 0.743 |
| HYPERION | 581 | 0.489 | 50 | 0.943 | 0.3 | 2,000 | 0.913 |

**eTable 7** – Power simulations for HYPERION dataset with neurologic outcome.

| **Dataset** | **N** | **TTM prevalence** | **Responsive subgroup (%)** | **Baseline risk** | **Assumed absolute treatment effect** | **Simulations** | **Power** |
| --- | --- | --- | --- | --- | --- | --- | --- |
| eICU | 2,082 | 0.303 | 20 | 0.484 | 0.1 | 2,000 | 0.402 |
| eICU | 2,082 | 0.303 | 30 | 0.484 | 0.1 | 2,000 | 0.511 |
| eICU | 2,082 | 0.303 | 50 | 0.484 | 0.1 | 2,000 | 0.582 |
| eICU | 2,082 | 0.303 | 20 | 0.484 | 0.2 | 2,000 | 0.934 |
| eICU | 2,082 | 0.303 | 30 | 0.484 | 0.2 | 2,000 | 0.974 |
| eICU | 2,082 | 0.303 | 50 | 0.484 | 0.2 | 2,000 | 0.99 |
| eICU | 2,082 | 0.303 | 20 | 0.484 | 0.3 | 2,000 | 1 |
| eICU | 2,082 | 0.303 | 30 | 0.484 | 0.3 | 2,000 | 1 |
| eICU | 2,082 | 0.303 | 50 | 0.484 | 0.3 | 2,000 | 1 |

**eTable 8** – Power simulations for eICU dataset with neurologic outcome.

| **Dataset** | **N** | **TTM prevalence** | **Responsive subgroup (%)** | **Baseline risk** | **Assumed absolute treatment effect** | **Simulations** | **Power** |
| --- | --- | --- | --- | --- | --- | --- | --- |
| MIMIC-IV | 552 | 0.504 | 20 | 0.664 | 0.1 | 2,000 | 0.194 |
| MIMIC-IV | 552 | 0.504 | 30 | 0.664 | 0.1 | 2,000 | 0.228 |
| MIMIC-IV | 552 | 0.504 | 50 | 0.664 | 0.1 | 2,000 | 0.259 |
| MIMIC-IV | 552 | 0.504 | 20 | 0.664 | 0.2 | 2,000 | 0.644 |
| MIMIC-IV | 552 | 0.504 | 30 | 0.664 | 0.2 | 2,000 | 0.758 |
| MIMIC-IV | 552 | 0.504 | 50 | 0.664 | 0.2 | 2,000 | 0.846 |
| MIMIC-IV | 552 | 0.504 | 20 | 0.664 | 0.3 | 2,000 | 0.988 |
| MIMIC-IV | 552 | 0.504 | 30 | 0.664 | 0.3 | 2,000 | 0.998 |
| MIMIC-IV | 552 | 0.504 | 50 | 0.664 | 0.3 | 2,000 | 1 |

**eTable 9** – Power simulations for MIMIC-IV dataset with neurologic outcome.

| **Dataset** | **N** | **TTM prevalence** | **Responsive subgroup (%)** | **Baseline risk** | **Assumed absolute treatment effect** | **Simulations** | **Power** |
| --- | --- | --- | --- | --- | --- | --- | --- |
| PMAP | 1,292 | 0.481 | 20 | 0.414 | 0.1 | 2,000 | 0.294 |
| PMAP | 1,292 | 0.481 | 30 | 0.414 | 0.1 | 2,000 | 0.374 |
| PMAP | 1,292 | 0.481 | 50 | 0.414 | 0.1 | 2,000 | 0.414 |
| PMAP | 1,292 | 0.481 | 20 | 0.414 | 0.2 | 2,000 | 0.824 |
| PMAP | 1,292 | 0.481 | 30 | 0.414 | 0.2 | 2,000 | 0.908 |
| PMAP | 1,292 | 0.481 | 50 | 0.414 | 0.2 | 2,000 | 0.952 |
| PMAP | 1,292 | 0.481 | 20 | 0.414 | 0.3 | 2,000 | 0.994 |
| PMAP | 1,292 | 0.481 | 30 | 0.414 | 0.3 | 2,000 | 1 |
| PMAP | 1,292 | 0.481 | 50 | 0.414 | 0.3 | 2,000 | 1 |

**eTable 10** – Power simulations for PMAP dataset with neurologic outcome.

| **Dataset** | **N** | **TTM prevalence** | **Responsive subgroup (%)** | **Baseline risk** | **Assumed absolute treatment effect** | **Simulations** | **Power** |
| --- | --- | --- | --- | --- | --- | --- | --- |
| HYPERION | 581 | 0.489 | 20 | 0.832 | 0.1 | 2,000 | 0.359 |
| HYPERION | 581 | 0.489 | 30 | 0.832 | 0.1 | 2,000 | 0.444 |
| HYPERION | 581 | 0.489 | 50 | 0.832 | 0.1 | 2,000 | 0.538 |
| HYPERION | 581 | 0.489 | 20 | 0.832 | 0.2 | 2,000 | 0.993 |
| HYPERION | 581 | 0.489 | 30 | 0.832 | 0.2 | 2,000 | 0.999 |
| HYPERION | 581 | 0.489 | 50 | 0.832 | 0.2 | 2,000 | 1 |
| HYPERION | 581 | 0.489 | 20 | 0.832 | 0.3 | 2,000 | 0.989 |
| HYPERION | 581 | 0.489 | 30 | 0.832 | 0.3 | 2,000 | 0.999 |
| HYPERION | 581 | 0.489 | 50 | 0.832 | 0.3 | 2,000 | 0.962 |

**eTable 11** – Power simulations for HYPERION dataset with hospital mortality outcome.

| **Dataset** | **N** | **TTM prevalence** | **Responsive subgroup (%)** | **Baseline risk** | **Assumed absolute treatment effect** | **Simulations** | **Power** |
| --- | --- | --- | --- | --- | --- | --- | --- |
| eICU | 2,082 | 0.303 | 20 | 0.516 | 0.1 | 2,000 | 0.404 |
| eICU | 2,082 | 0.303 | 30 | 0.516 | 0.1 | 2,000 | 0.485 |
| eICU | 2,082 | 0.303 | 50 | 0.516 | 0.1 | 2,000 | 0.552 |
| eICU | 2,082 | 0.303 | 20 | 0.516 | 0.2 | 2,000 | 0.932 |
| eICU | 2,082 | 0.303 | 30 | 0.516 | 0.2 | 2,000 | 0.974 |
| eICU | 2,082 | 0.303 | 50 | 0.516 | 0.2 | 2,000 | 0.992 |
| eICU | 2,082 | 0.303 | 20 | 0.516 | 0.3 | 2,000 | 1 |
| eICU | 2,082 | 0.303 | 30 | 0.516 | 0.3 | 2,000 | 1 |
| eICU | 2,082 | 0.303 | 50 | 0.516 | 0.3 | 2,000 | 1 |

**eTable 12** – Power simulations for eICU dataset with hospital mortality outcome.

| **Dataset** | **N** | **TTM prevalence** | **Responsive subgroup (%)** | **Baseline risk** | **Assumed absolute treatment effect** | **Simulations** | **Power** |
| --- | --- | --- | --- | --- | --- | --- | --- |
| MIMIC-IV | 552 | 0.504 | 20 | 0.336 | 0.1 | 2,000 | 0.162 |
| MIMIC-IV | 552 | 0.504 | 30 | 0.336 | 0.1 | 2,000 | 0.195 |
| MIMIC-IV | 552 | 0.504 | 50 | 0.336 | 0.1 | 2,000 | 0.241 |
| MIMIC-IV | 552 | 0.504 | 20 | 0.336 | 0.2 | 2,000 | 0.472 |
| MIMIC-IV | 552 | 0.504 | 30 | 0.336 | 0.2 | 2,000 | 0.584 |
| MIMIC-IV | 552 | 0.504 | 50 | 0.336 | 0.2 | 2,000 | 0.662 |
| MIMIC-IV | 552 | 0.504 | 20 | 0.336 | 0.3 | 2,000 | 0.813 |
| MIMIC-IV | 552 | 0.504 | 30 | 0.336 | 0.3 | 2,000 | 0.908 |
| MIMIC-IV | 552 | 0.504 | 50 | 0.336 | 0.3 | 2,000 | 0.94 |

**eTable 13** – Power simulations for MIMIC-IV dataset with hospital mortality outcome.

| **Dataset** | **N** | **TTM prevalence** | **Responsive subgroup (%)** | **Baseline risk** | **Assumed absolute treatment effect** | **Simulations** | **Power** |
| --- | --- | --- | --- | --- | --- | --- | --- |
| PMAP | 1,292 | 0.481 | 20 | 0.414 | 0.1 | 2,000 | 0.294 |
| PMAP | 1,292 | 0.481 | 30 | 0.414 | 0.1 | 2,000 | 0.374 |
| PMAP | 1,292 | 0.481 | 50 | 0.414 | 0.1 | 2,000 | 0.414 |
| PMAP | 1,292 | 0.481 | 20 | 0.414 | 0.2 | 2,000 | 0.824 |
| PMAP | 1,292 | 0.481 | 30 | 0.414 | 0.2 | 2,000 | 0.908 |
| PMAP | 1,292 | 0.481 | 50 | 0.414 | 0.2 | 2,000 | 0.952 |
| PMAP | 1,292 | 0.481 | 20 | 0.414 | 0.3 | 2,000 | 0.994 |
| PMAP | 1,292 | 0.481 | 30 | 0.414 | 0.3 | 2,000 | 1 |
| PMAP | 1,292 | 0.481 | 50 | 0.414 | 0.3 | 2,000 | 1 |

**eTable 14** – Power simulations for PMAP dataset with hospital mortality outcome.

### S.11 GATES Numeric Results

| **Dataset** | **Quintile** | **n** | **Mean Cate** | **GATE** | **F p-value** | **Spearman ρ** | **Spearman p** | **95% CI** |
| --- | --- | --- | --- | --- | --- | --- | --- | --- |
| eICU | 1 | 111 | 0.019 | -0.062 | 0.0971 | 0.4 | 0.5046 | [-0.242, 0.118] |
| eICU | 2 | 110 | 0.069 | 0.007 | 0.0971 | 0.4 | 0.5046 | [-0.183, 0.197] |
| eICU | 3 | 111 | 0.099 | 0.11 | 0.0971 | 0.4 | 0.5046 | [-0.073, 0.293] |
| eICU | 4 | 110 | 0.132 | -0.143 | 0.0971 | 0.4 | 0.5046 | [-0.332, 0.046] |
| eICU | 5 | 111 | 0.178 | 0.208 | 0.0971 | 0.4 | 0.5046 | [0.008, 0.408] |
| HYPERION | 1 | 35 | -0.042 | 0.124 | 0.1332 | -0.9 | 0.0374 | [-0.026, 0.274] |
| HYPERION | 2 | 35 | -0.019 | 0.023 | 0.1332 | -0.9 | 0.0374 | [-0.219, 0.264] |
| HYPERION | 3 | 35 | 0.003 | 0.043 | 0.1332 | -0.9 | 0.0374 | [-0.217, 0.302] |
| HYPERION | 4 | 35 | 0.026 | -0.02 | 0.1332 | -0.9 | 0.0374 | [-0.352, 0.312] |
| HYPERION | 5 | 35 | 0.063 | -0.339 | 0.1332 | -0.9 | 0.0374 | [-0.667, -0.011] |
| MIMIC-IV | 1 | 34 | -0.027 | -0.09 | 0.5104 | 0.8 | 0.1041 | [-0.361, 0.181] |
| MIMIC-IV | 2 | 33 | 0.038 | -0.177 | 0.5104 | 0.8 | 0.1041 | [-0.539, 0.185] |
| MIMIC-IV | 3 | 34 | 0.069 | 0.02 | 0.5104 | 0.8 | 0.1041 | [-0.276, 0.315] |
| MIMIC-IV | 4 | 33 | 0.096 | 0.226 | 0.5104 | 0.8 | 0.1041 | [-0.118, 0.57] |
| MIMIC-IV | 5 | 34 | 0.138 | 0.024 | 0.5104 | 0.8 | 0.1041 | [-0.337, 0.384] |
| PMAP | 1 | 85 | -0.013 | 0.168 | 0.3184 | 0.3 | 0.6238 | [-0.032, 0.368] |
| PMAP | 2 | 85 | 0.024 | 0.058 | 0.3184 | 0.3 | 0.6238 | [-0.14, 0.255] |
| PMAP | 3 | 84 | 0.045 | -0.069 | 0.3184 | 0.3 | 0.6238 | [-0.274, 0.135] |
| PMAP | 4 | 85 | 0.069 | 0.131 | 0.3184 | 0.3 | 0.6238 | [-0.075, 0.338] |
| PMAP | 5 | 85 | 0.111 | 0.222 | 0.3184 | 0.3 | 0.6238 | [0.014, 0.43] |

**eTable 15 –** GATES results for hospital mortality. IPW-weighted group average treatment effects within quintiles of predicted CATE from CausalForestDML. Unweighted OLS was used for HYPERION. F-statistic and p-value test joint significance of quintile–treatment interactions. Spearman ρ tests monotonicity of GATE estimates across quintiles.

### S.12 Feature List and Per-Dataset Missing Data Report

| **Variable** | **Missing, n** | **Missing, %** | **Missing in No-TTM Group, %** | **Missing in TTM Group, %** | **Absolute Difference, %** | **P Value** | **Statistical Test** |
| --- | --- | --- | --- | --- | --- | --- | --- |
| nurse_first_QTc | 1837 | 99.7285559 | 99.7568882 | 99.6710526 | 0.08583554 | 0.66739245 | Fisher |
| lab_first_troponin - T | 1531 | 83.1161781 | 83.7925446 | 81.7434211 | 2.04912352 | 0.26960917 | Chi-square |
| lab_first_Respiratory Rate | 1497 | 81.2703583 | 81.280389 | 81.25 | 0.03038898 | 0.98745785 | Chi-square |
| lab_first_troponin - I | 947 | 51.4115092 | 54.1329011 | 45.8881579 | 8.24474324 | 0.00087084 | Chi-square |
| lab_first_lactate | 559 | 30.3474484 | 35.0081037 | 20.8881579 | 14.1199458 | 5.71E-10 | Chi-square |
| lab_first_magnesium | 529 | 28.7187839 | 34.1166937 | 17.7631579 | 16.3535358 | 2.99E-13 | Chi-square |
| lab_first_FiO2 | 389 | 21.1183496 | 21.9611021 | 19.4078947 | 2.55320737 | 0.20676759 | Chi-square |
| nurse_first_GCS Total | 352 | 19.1096634 | 17.0178282 | 23.3552632 | 6.33743496 | 0.00114136 | Chi-square |
| lab_first_pH | 320 | 17.3724213 | 19.286872 | 13.4868421 | 5.80002986 | 0.00200427 | Chi-square |
| nurse_first_Motor | 310 | 16.8295331 | 15.3970827 | 19.7368421 | 4.33975945 | 0.01922989 | Chi-square |
| lab_first_paO2 | 294 | 15.9609121 | 17.9092382 | 12.0065789 | 5.9026593 | 0.00114318 | Chi-square |
| lab_first_paCO2 | 283 | 15.3637351 | 17.3419773 | 11.3486842 | 5.9932931 | 0.00079563 | Chi-square |
| lab_first_bicarbonate | 221 | 11.9978284 | 16.4505673 | 2.96052632 | 13.4900409 | 5.35E-17 | Chi-square |
| lab_first_calcium | 193 | 10.4777416 | 13.2090762 | 4.93421053 | 8.27486565 | 4.96E-08 | Chi-square |
| lab_first_WBC x 1000 | 159 | 8.63192182 | 9.96758509 | 5.92105263 | 4.04653246 | 0.00363731 | Chi-square |
| lab_first_platelets x 1000 | 158 | 8.57763301 | 10.1296596 | 5.42763158 | 4.70202806 | 0.00070211 | Chi-square |
| nurse_first_O2 Saturation | 154 | 8.36047774 | 9.07617504 | 6.90789474 | 2.1682803 | 0.1138854 | Chi-square |
| lab_first_Hgb | 146 | 7.92616721 | 9.31928687 | 5.09868421 | 4.22060266 | 0.0016154 | Chi-square |
| nurse_first_Non-Invasive BP Mean | 136 | 7.38327904 | 8.99513776 | 4.11184211 | 4.88329566 | 0.000164 | Chi-square |
| diagnosis_initial rhythm: asystole | 127 | 6.8946797 | 5.75364668 | 9.21052632 | 3.45687964 | 0.00589396 | Chi-square |
| diagnosis_initial rhythm: pulseless electrical activity | 127 | 6.8946797 | 5.75364668 | 9.21052632 | 3.45687964 | 0.00589396 | Chi-square |
| diagnosis_initial rhythm: ventricular fibrillation | 127 | 6.8946797 | 5.75364668 | 9.21052632 | 3.45687964 | 0.00589396 | Chi-square |
| diagnosis_initial rhythm: ventricular tachycardia | 127 | 6.8946797 | 5.75364668 | 9.21052632 | 3.45687964 | 0.00589396 | Chi-square |
| diagnosis_ventricular fibrillation | 127 | 6.8946797 | 5.75364668 | 9.21052632 | 3.45687964 | 0.00589396 | Chi-square |
| diagnosis_ventricular tachycardia | 127 | 6.8946797 | 5.75364668 | 9.21052632 | 3.45687964 | 0.00589396 | Chi-square |
| lab_first_glucose | 120 | 6.51465798 | 8.75202593 | 1.97368421 | 6.77834172 | 2.97E-08 | Chi-square |
| lab_first_BUN | 117 | 6.35179153 | 8.34683955 | 2.30263158 | 6.04420797 | 5.69E-07 | Chi-square |
| lab_first_creatinine | 115 | 6.2432139 | 8.26580227 | 2.13815789 | 6.12764437 | 3.20E-07 | Chi-square |
| lab_first_sodium | 104 | 5.64603692 | 7.61750405 | 1.64473684 | 5.97276721 | 1.76E-07 | Chi-square |
| nurse_first_Non-Invasive BP Diastolic | 97 | 5.2660152 | 6.32090762 | 3.125 | 3.19590762 | 0.00387965 | Chi-square |
| nurse_first_Non-Invasive BP Systolic | 97 | 5.2660152 | 6.32090762 | 3.125 | 3.19590762 | 0.00387965 | Chi-square |
| lab_first_potassium | 96 | 5.21172638 | 7.13128039 | 1.31578947 | 5.81549092 | 1.29E-07 | Chi-square |
| bmi | 61 | 3.31161781 | 4.6191248 | 0.65789474 | 3.96123006 | 7.91E-06 | Chi-square |
| nurse_first_Heart Rate | 52 | 2.82301846 | 4.21393841 | 0 | 4.21393841 | 2.83E-07 | Chi-square |
| age | 0 | 0 | 0 | 0 | 0 |  | Failed |
| gender | 0 | 0 | 0 | 0 | 0 |  | Failed |

**eTable 16** – Complete feature list and missing variable report for eICU-CRD dataset.

| **Variable** | **Missing, n** | **Missing, %** | **Missing in No-TTM Group, %** | **Missing in TTM Group, %** | **Absolute Difference, %** | **P Value** | **Statistical Test** |
| --- | --- | --- | --- | --- | --- | --- | --- |
| lab_first_ldl_measured | 611 | 100 | 100 | 100 | 0 |  | Failed |
| chart_first_fio2_(ch) | 610 | 99.8363339 | 99.6978852 | 100 | 0.3021148 | 1 | Fisher |
| chart_first_flow_(ch) | 610 | 99.8363339 | 99.6978852 | 100 | 0.3021148 | 1 | Fisher |
| chart_first_intra_cranial_pressure_#2 | 610 | 99.8363339 | 100 | 99.6428571 | 0.35714286 | 0.45826514 | Fisher |
| chart_first_qtc | 610 | 99.8363339 | 100 | 99.6428571 | 0.35714286 | 0.45826514 | Fisher |
| chart_first_vd/vt_ratio | 610 | 99.8363339 | 100 | 99.6428571 | 0.35714286 | 0.45826514 | Fisher |
| output_first_qtc | 610 | 99.8363339 | 100 | 99.6428571 | 0.35714286 | 0.45826514 | Fisher |
| chart_first_total_protein | 609 | 99.6726678 | 99.6978852 | 99.6428571 | 0.05502805 | 1 | Fisher |
| output_first_total_protein | 609 | 99.6726678 | 99.6978852 | 99.6428571 | 0.05502805 | 1 | Fisher |
| chart_first_feeding_weight | 608 | 99.5090016 | 99.3957704 | 99.6428571 | 0.24708675 | 1 | Fisher |
| chart_first_height | 608 | 99.5090016 | 99.6978852 | 99.2857143 | 0.41217091 | 0.59576596 | Fisher |
| chart_first_height_(cm) | 608 | 99.5090016 | 99.6978852 | 99.2857143 | 0.41217091 | 0.59576596 | Fisher |
| chart_first_intra_cranial_pressure | 608 | 99.5090016 | 99.3957704 | 99.6428571 | 0.24708675 | 1 | Fisher |
| chart_first_spont_vt | 608 | 99.5090016 | 99.3957704 | 99.6428571 | 0.24708675 | 1 | Fisher |
| lab_first_digoxin | 608 | 99.5090016 | 99.3957704 | 99.6428571 | 0.24708675 | 1 | Fisher |
| chart_first_cardiac_index_(ci_nicom) | 606 | 99.1816694 | 98.489426 | 100 | 1.51057402 | 0.06583335 | Fisher |
| chart_first_d-dimer | 606 | 99.1816694 | 98.7915408 | 99.6428571 | 0.85131636 | 0.38168149 | Fisher |
| chart_first_fio2_(ecmo) | 606 | 99.1816694 | 98.489426 | 100 | 1.51057402 | 0.06583335 | Fisher |
| chart_first_flow_(ecmo) | 606 | 99.1816694 | 98.489426 | 100 | 1.51057402 | 0.06583335 | Fisher |
| chart_first_peco2 | 606 | 99.1816694 | 99.3957704 | 98.9285714 | 0.46719896 | 0.66486525 | Fisher |
| output_first_d-dimer | 606 | 99.1816694 | 98.7915408 | 99.6428571 | 0.85131636 | 0.38168149 | Fisher |
| chart_first_impella_catheter_position | 603 | 98.690671 | 97.5830816 | 100 | 2.41691843 | 0.00897398 | Fisher |
| chart_first_gi_ph | 602 | 98.5270049 | 99.3957704 | 97.5 | 1.89577039 | 0.08753647 | Fisher |
| chart_first_manual_blood_pressure_diastolic_left | 600 | 98.1996727 | 99.6978852 | 96.4285714 | 3.26931377 | 0.00245937 | Chi-square |
| chart_first_access_pressure | 594 | 97.2176759 | 96.0725076 | 98.5714286 | 2.49892102 | 0.0613005 | Chi-square |
| chart_first_blood_flow_(ml/min) | 594 | 97.2176759 | 96.0725076 | 98.5714286 | 2.49892102 | 0.0613005 | Chi-square |
| lab_first_brain_natiuretic_peptide_(bnp) | 591 | 96.7266776 | 97.2809668 | 96.0714286 | 1.2095382 | 0.40248531 | Chi-square |
| chart_first_svo2 | 589 | 96.3993453 | 94.2598187 | 98.9285714 | 4.6687527 | 0.00202629 | Chi-square |
| chart_first_lipase | 574 | 93.9443535 | 92.1450151 | 96.0714286 | 3.92641347 | 0.04261513 | Chi-square |
| output_first_lipase | 574 | 93.9443535 | 92.1450151 | 96.0714286 | 3.92641347 | 0.04261513 | Chi-square |
| chart_first_pulmonary_artery_pressure_mean | 570 | 93.289689 | 89.1238671 | 98.2142857 | 9.09041864 | 7.65E-06 | Chi-square |
| chart_first_bis_index_range | 530 | 86.7430442 | 95.1661631 | 76.7857143 | 18.3804489 | 2.46E-11 | Chi-square |
| chart_first_hematocrit_(whole_blood_-_calc) | 515 | 84.2880524 | 81.570997 | 87.5 | 5.92900302 | 0.04479461 | Chi-square |
| chart_first_daily_weight | 512 | 83.797054 | 86.4048338 | 80.7142857 | 5.69054812 | 0.05716851 | Chi-square |
| chart_first_absolute_count_-_eos | 500 | 81.8330606 | 84.8942598 | 78.2142857 | 6.6799741 | 0.03286391 | Chi-square |
| chart_first_etco2 | 452 | 73.9770867 | 80.6646526 | 66.0714286 | 14.593224 | 4.20E-05 | Chi-square |
| chart_first_glucose_(whole_blood) | 436 | 71.3584288 | 64.652568 | 79.2857143 | 14.6331463 | 6.71E-05 | Chi-square |
| chart_first_ph_(venous) | 420 | 68.7397709 | 74.3202417 | 62.1428571 | 12.1773845 | 0.00121478 | Chi-square |
| chart_first_arterial_blood_pressure_alarm_-_high | 412 | 67.4304419 | 66.4652568 | 68.5714286 | 2.10617177 | 0.57990831 | Chi-square |
| chart_first_arterial_blood_pressure_mean | 275 | 45.0081833 | 39.8791541 | 51.0714286 | 11.1922745 | 0.00559288 | Chi-square |
| output_first_alt | 236 | 38.6252046 | 45.3172205 | 30.7142857 | 14.6029348 | 0.00022086 | Chi-square |
| lab_first_troponin-t | 227 | 37.1522095 | 48.6404834 | 23.5714286 | 25.0690548 | 1.66E-10 | Chi-square |
| chart_20_g_infiltration_scale_Grade 0 | 195 | 31.9148936 | 32.9305136 | 30.7142857 | 2.21622788 | 0.55817789 | Chi-square |
| chart_20_g_infiltration_scale_Grade 2 | 195 | 31.9148936 | 32.9305136 | 30.7142857 | 2.21622788 | 0.55817789 | Chi-square |
| chart_20_gauge_placed_in_the_field | 195 | 31.9148936 | 32.9305136 | 30.7142857 | 2.21622788 | 0.55817789 | Chi-square |
| chart_alarms_on | 195 | 31.9148936 | 32.9305136 | 30.7142857 | 2.21622788 | 0.55817789 | Chi-square |
| chart_angio_dressing_#_1_Bandaid | 195 | 31.9148936 | 32.9305136 | 30.7142857 | 2.21622788 | 0.55817789 | Chi-square |
| chart_angio_dressing_#_2_Bandaid | 195 | 31.9148936 | 32.9305136 | 30.7142857 | 2.21622788 | 0.55817789 | Chi-square |
| chart_angio_site_#_1_R Femoral | 195 | 31.9148936 | 32.9305136 | 30.7142857 | 2.21622788 | 0.55817789 | Chi-square |
| chart_art_bubble_on_(ch) | 195 | 31.9148936 | 32.9305136 | 30.7142857 | 2.21622788 | 0.55817789 | Chi-square |
| chart_back_care | 195 | 31.9148936 | 32.9305136 | 30.7142857 | 2.21622788 | 0.55817789 | Chi-square |
| chart_cardiac_assist_cannula_site_appear_WNL | 195 | 31.9148936 | 32.9305136 | 30.7142857 | 2.21622788 | 0.55817789 | Chi-square |
| chart_cervical_collar_status_On | 195 | 31.9148936 | 32.9305136 | 30.7142857 | 2.21622788 | 0.55817789 | Chi-square |
| chart_circuit_configuration_(ch)_VA | 195 | 31.9148936 | 32.9305136 | 30.7142857 | 2.21622788 | 0.55817789 | Chi-square |
| chart_corneal_reflex_left_Absent | 195 | 31.9148936 | 32.9305136 | 30.7142857 | 2.21622788 | 0.55817789 | Chi-square |
| chart_corneal_reflex_left_Intact | 195 | 31.9148936 | 32.9305136 | 30.7142857 | 2.21622788 | 0.55817789 | Chi-square |
| chart_cv_-_past_medical_history_CAD | 195 | 31.9148936 | 32.9305136 | 30.7142857 | 2.21622788 | 0.55817789 | Chi-square |
| chart_cv_-_past_medical_history_Hypertension | 195 | 31.9148936 | 32.9305136 | 30.7142857 | 2.21622788 | 0.55817789 | Chi-square |
| chart_daily_wake_up_No, at goal RASS | 195 | 31.9148936 | 32.9305136 | 30.7142857 | 2.21622788 | 0.55817789 | Chi-square |
| chart_daily_wake_up_Yes, sedation adjusted or stopped | 195 | 31.9148936 | 32.9305136 | 30.7142857 | 2.21622788 | 0.55817789 | Chi-square |
| chart_daily_wake_up_deferred_Neuromusc Block | 195 | 31.9148936 | 32.9305136 | 30.7142857 | 2.21622788 | 0.55817789 | Chi-square |
| chart_education_topic_Cardioversion | 195 | 31.9148936 | 32.9305136 | 30.7142857 | 2.21622788 | 0.55817789 | Chi-square |
| chart_emotional_/_physical_/_sexual_harm_by_partner_or_close_relation | 195 | 31.9148936 | 32.9305136 | 30.7142857 | 2.21622788 | 0.55817789 | Chi-square |
| chart_ett_location_Oral-R | 195 | 31.9148936 | 32.9305136 | 30.7142857 | 2.21622788 | 0.55817789 | Chi-square |
| chart_eye_care | 195 | 31.9148936 | 32.9305136 | 30.7142857 | 2.21622788 | 0.55817789 | Chi-square |
| chart_gag_reflex_Absent | 195 | 31.9148936 | 32.9305136 | 30.7142857 | 2.21622788 | 0.55817789 | Chi-square |
| chart_gag_reflex_Intact | 195 | 31.9148936 | 32.9305136 | 30.7142857 | 2.21622788 | 0.55817789 | Chi-square |
| chart_gcs_-_eye_opening_To Pain | 195 | 31.9148936 | 32.9305136 | 30.7142857 | 2.21622788 | 0.55817789 | Chi-square |
| chart_gcs_-_verbal_response_No Response | 195 | 31.9148936 | 32.9305136 | 30.7142857 | 2.21622788 | 0.55817789 | Chi-square |
| chart_gcs_-_verbal_response_Oriented | 195 | 31.9148936 | 32.9305136 | 30.7142857 | 2.21622788 | 0.55817789 | Chi-square |
| chart_goal_richmond-ras_scale_ No eye contact | 195 | 31.9148936 | 32.9305136 | 30.7142857 | 2.21622788 | 0.55817789 | Chi-square |
| chart_heart_rhythm_2nd AV M2 (Second degree AV Block - Mobitz 2) | 195 | 31.9148936 | 32.9305136 | 30.7142857 | 2.21622788 | 0.55817789 | Chi-square |
| chart_heart_rhythm_3rd AV (Complete Heart Block) | 195 | 31.9148936 | 32.9305136 | 30.7142857 | 2.21622788 | 0.55817789 | Chi-square |
| chart_heart_rhythm_AF (Atrial Fibrillation) | 195 | 31.9148936 | 32.9305136 | 30.7142857 | 2.21622788 | 0.55817789 | Chi-square |
| chart_heart_rhythm_SR (Sinus Rhythm) | 195 | 31.9148936 | 32.9305136 | 30.7142857 | 2.21622788 | 0.55817789 | Chi-square |
| chart_heart_rhythm_ST (Sinus Tachycardia) | 195 | 31.9148936 | 32.9305136 | 30.7142857 | 2.21622788 | 0.55817789 | Chi-square |
| chart_heart_rhythm_V Paced | 195 | 31.9148936 | 32.9305136 | 30.7142857 | 2.21622788 | 0.55817789 | Chi-square |
| chart_humidification_Active | 195 | 31.9148936 | 32.9305136 | 30.7142857 | 2.21622788 | 0.55817789 | Chi-square |
| chart_impaired_fluid_balance_ncp_-_interventions_Monitoring electrolytes / renal function | 195 | 31.9148936 | 32.9305136 | 30.7142857 | 2.21622788 | 0.55817789 | Chi-square |
| chart_impaired_skin_cleanse_#6_Foam Cleanser | 195 | 31.9148936 | 32.9305136 | 30.7142857 | 2.21622788 | 0.55817789 | Chi-square |
| chart_incision_#1-_location_Chin | 195 | 31.9148936 | 32.9305136 | 30.7142857 | 2.21622788 | 0.55817789 | Chi-square |
| chart_incision_#1-_location_Tracheostomy | 195 | 31.9148936 | 32.9305136 | 30.7142857 | 2.21622788 | 0.55817789 | Chi-square |
| chart_incision_#1-_treatment_Ace Wrap | 195 | 31.9148936 | 32.9305136 | 30.7142857 | 2.21622788 | 0.55817789 | Chi-square |
| chart_incision_#1-_treatment_Xeroform | 195 | 31.9148936 | 32.9305136 | 30.7142857 | 2.21622788 | 0.55817789 | Chi-square |
| chart_intra_aortic_ballon_pump_setting_1:1 | 195 | 31.9148936 | 32.9305136 | 30.7142857 | 2.21622788 | 0.55817789 | Chi-square |
| chart_intra_aortic_balloon_pump_setting_1:2 | 195 | 31.9148936 | 32.9305136 | 30.7142857 | 2.21622788 | 0.55817789 | Chi-square |
| chart_is_the_spokesperson_the_health_care_proxy | 195 | 31.9148936 | 32.9305136 | 30.7142857 | 2.21622788 | 0.55817789 | Chi-square |
| chart_lue_color_Normal | 195 | 31.9148936 | 32.9305136 | 30.7142857 | 2.21622788 | 0.55817789 | Chi-square |
| chart_motor_l_arm_Localizes | 195 | 31.9148936 | 32.9305136 | 30.7142857 | 2.21622788 | 0.55817789 | Chi-square |
| chart_motor_l_leg_Localizes | 195 | 31.9148936 | 32.9305136 | 30.7142857 | 2.21622788 | 0.55817789 | Chi-square |
| chart_neuro_drain_#1_level_15 cm | 195 | 31.9148936 | 32.9305136 | 30.7142857 | 2.21622788 | 0.55817789 | Chi-square |
| chart_neuro_drain_landmark_Tragus | 195 | 31.9148936 | 32.9305136 | 30.7142857 | 2.21622788 | 0.55817789 | Chi-square |
| chart_pain_level_Unable to Score | 195 | 31.9148936 | 32.9305136 | 30.7142857 | 2.21622788 | 0.55817789 | Chi-square |
| chart_pain_level_response_Unable to Score | 195 | 31.9148936 | 32.9305136 | 30.7142857 | 2.21622788 | 0.55817789 | Chi-square |
| chart_pain_management_Repositioned | 195 | 31.9148936 | 32.9305136 | 30.7142857 | 2.21622788 | 0.55817789 | Chi-square |
| chart_par-circulation_BP +/- 20% of pre-anesthesthetic level | 195 | 31.9148936 | 32.9305136 | 30.7142857 | 2.21622788 | 0.55817789 | Chi-square |
| chart_past_medical_history_COPD | 195 | 31.9148936 | 32.9305136 | 30.7142857 | 2.21622788 | 0.55817789 | Chi-square |
| chart_patient/family_informed_----- | 195 | 31.9148936 | 32.9305136 | 30.7142857 | 2.21622788 | 0.55817789 | Chi-square |
| chart_performance_level_(r)_P6 | 195 | 31.9148936 | 32.9305136 | 30.7142857 | 2.21622788 | 0.55817789 | Chi-square |
| chart_performance_level_Auto | 195 | 31.9148936 | 32.9305136 | 30.7142857 | 2.21622788 | 0.55817789 | Chi-square |
| chart_performance_level_P2 | 195 | 31.9148936 | 32.9305136 | 30.7142857 | 2.21622788 | 0.55817789 | Chi-square |
| chart_post-op_care_ncp_-_interventions_Assess surgical wounds and drains | 195 | 31.9148936 | 32.9305136 | 30.7142857 | 2.21622788 | 0.55817789 | Chi-square |
| chart_problem_list_.H/O abdominal compartment syndrome / Intraabdominal Hypertension (IAH, ACS) | 195 | 31.9148936 | 32.9305136 | 30.7142857 | 2.21622788 | 0.55817789 | Chi-square |
| chart_problem_list_.H/O anemia, hemolytic | 195 | 31.9148936 | 32.9305136 | 30.7142857 | 2.21622788 | 0.55817789 | Chi-square |
| chart_problem_list_.H/O atrial fibrillation (Afib) | 195 | 31.9148936 | 32.9305136 | 30.7142857 | 2.21622788 | 0.55817789 | Chi-square |
| chart_problem_list_.H/O tobacco¬†use, current | 195 | 31.9148936 | 32.9305136 | 30.7142857 | 2.21622788 | 0.55817789 | Chi-square |
| chart_problem_list_Cardiac arrest | 195 | 31.9148936 | 32.9305136 | 30.7142857 | 2.21622788 | 0.55817789 | Chi-square |
| chart_problem_list_Cerebrovascular disease, other | 195 | 31.9148936 | 32.9305136 | 30.7142857 | 2.21622788 | 0.55817789 | Chi-square |
| chart_problem_list_Myocardial infarction, acute (AMI, STEMI, NSTEMI) | 195 | 31.9148936 | 32.9305136 | 30.7142857 | 2.21622788 | 0.55817789 | Chi-square |
| chart_problem_list_Pulmonary edema | 195 | 31.9148936 | 32.9305136 | 30.7142857 | 2.21622788 | 0.55817789 | Chi-square |
| chart_problem_list_Renal failure, acute (Acute renal failure, ARF, AKI) | 195 | 31.9148936 | 32.9305136 | 30.7142857 | 2.21622788 | 0.55817789 | Chi-square |
| chart_problem_list_Shock, cardiogenic | 195 | 31.9148936 | 32.9305136 | 30.7142857 | 2.21622788 | 0.55817789 | Chi-square |
| chart_problem_list_Tachycardia, Other | 195 | 31.9148936 | 32.9305136 | 30.7142857 | 2.21622788 | 0.55817789 | Chi-square |
| chart_problem_list_Ventricular tachycardia, sustained | 195 | 31.9148936 | 32.9305136 | 30.7142857 | 2.21622788 | 0.55817789 | Chi-square |
| chart_pupil_response_left_Non-reactive | 195 | 31.9148936 | 32.9305136 | 30.7142857 | 2.21622788 | 0.55817789 | Chi-square |
| chart_pupil_response_right_Non-reactive | 195 | 31.9148936 | 32.9305136 | 30.7142857 | 2.21622788 | 0.55817789 | Chi-square |
| chart_pupil_size_left_5mm | 195 | 31.9148936 | 32.9305136 | 30.7142857 | 2.21622788 | 0.55817789 | Chi-square |
| chart_pupil_size_left_Pinpoint | 195 | 31.9148936 | 32.9305136 | 30.7142857 | 2.21622788 | 0.55817789 | Chi-square |
| chart_pupil_size_right_Fully Dilated | 195 | 31.9148936 | 32.9305136 | 30.7142857 | 2.21622788 | 0.55817789 | Chi-square |
| chart_pupil_size_right_Pinpoint | 195 | 31.9148936 | 32.9305136 | 30.7142857 | 2.21622788 | 0.55817789 | Chi-square |
| chart_respiratory_effort_Normal | 195 | 31.9148936 | 32.9305136 | 30.7142857 | 2.21622788 | 0.55817789 | Chi-square |
| chart_rul_lung_sounds_Diminished | 195 | 31.9148936 | 32.9305136 | 30.7142857 | 2.21622788 | 0.55817789 | Chi-square |
| chart_seizure_duration_Status | 195 | 31.9148936 | 32.9305136 | 30.7142857 | 2.21622788 | 0.55817789 | Chi-square |
| chart_skin_condition_Diaphoretic | 195 | 31.9148936 | 32.9305136 | 30.7142857 | 2.21622788 | 0.55817789 | Chi-square |
| chart_slope_On | 195 | 31.9148936 | 32.9305136 | 30.7142857 | 2.21622788 | 0.55817789 | Chi-square |
| chart_speech_Normal | 195 | 31.9148936 | 32.9305136 | 30.7142857 | 2.21622788 | 0.55817789 | Chi-square |
| chart_st_segment_monitoring_on | 195 | 31.9148936 | 32.9305136 | 30.7142857 | 2.21622788 | 0.55817789 | Chi-square |
| chart_stool_consistency_Loose | 195 | 31.9148936 | 32.9305136 | 30.7142857 | 2.21622788 | 0.55817789 | Chi-square |
| chart_stroke_ncp_-_type_Hemorrhagic | 195 | 31.9148936 | 32.9305136 | 30.7142857 | 2.21622788 | 0.55817789 | Chi-square |
| chart_svo2_sqi_1 | 195 | 31.9148936 | 32.9305136 | 30.7142857 | 2.21622788 | 0.55817789 | Chi-square |
| chart_tobacco_use_history_Never used | 195 | 31.9148936 | 32.9305136 | 30.7142857 | 2.21622788 | 0.55817789 | Chi-square |
| chart_urine_appearance_Cloudy | 195 | 31.9148936 | 32.9305136 | 30.7142857 | 2.21622788 | 0.55817789 | Chi-square |
| input_amiodarone | 195 | 31.9148936 | 32.9305136 | 30.7142857 | 2.21622788 | 0.55817789 | Chi-square |
| input_calcium_gluconate | 195 | 31.9148936 | 32.9305136 | 30.7142857 | 2.21622788 | 0.55817789 | Chi-square |
| input_diazepam_(valium) | 195 | 31.9148936 | 32.9305136 | 30.7142857 | 2.21622788 | 0.55817789 | Chi-square |
| input_diltiazem | 195 | 31.9148936 | 32.9305136 | 30.7142857 | 2.21622788 | 0.55817789 | Chi-square |
| input_famotidine_(pepcid) | 195 | 31.9148936 | 32.9305136 | 30.7142857 | 2.21622788 | 0.55817789 | Chi-square |
| input_heparin_sodium | 195 | 31.9148936 | 32.9305136 | 30.7142857 | 2.21622788 | 0.55817789 | Chi-square |
| input_lidocaine | 195 | 31.9148936 | 32.9305136 | 30.7142857 | 2.21622788 | 0.55817789 | Chi-square |
| input_lorazepam_(ativan) | 195 | 31.9148936 | 32.9305136 | 30.7142857 | 2.21622788 | 0.55817789 | Chi-square |
| input_magnesium_sulfate | 195 | 31.9148936 | 32.9305136 | 30.7142857 | 2.21622788 | 0.55817789 | Chi-square |
| input_norepinephrine | 195 | 31.9148936 | 32.9305136 | 30.7142857 | 2.21622788 | 0.55817789 | Chi-square |
| input_packed_red_blood_cells | 195 | 31.9148936 | 32.9305136 | 30.7142857 | 2.21622788 | 0.55817789 | Chi-square |
| input_po_intake | 195 | 31.9148936 | 32.9305136 | 30.7142857 | 2.21622788 | 0.55817789 | Chi-square |
| input_rocuronium | 195 | 31.9148936 | 32.9305136 | 30.7142857 | 2.21622788 | 0.55817789 | Chi-square |
| input_sodium_bicarbonate_8.4% | 195 | 31.9148936 | 32.9305136 | 30.7142857 | 2.21622788 | 0.55817789 | Chi-square |
| input_solution | 195 | 31.9148936 | 32.9305136 | 30.7142857 | 2.21622788 | 0.55817789 | Chi-square |
| input_tirofiban_(aggrastat) | 195 | 31.9148936 | 32.9305136 | 30.7142857 | 2.21622788 | 0.55817789 | Chi-square |
| input_vasopressin | 195 | 31.9148936 | 32.9305136 | 30.7142857 | 2.21622788 | 0.55817789 | Chi-square |
| med_alteplase | 195 | 31.9148936 | 32.9305136 | 30.7142857 | 2.21622788 | 0.55817789 | Chi-square |
| med_amiodarone | 195 | 31.9148936 | 32.9305136 | 30.7142857 | 2.21622788 | 0.55817789 | Chi-square |
| med_aspirin_ec | 195 | 31.9148936 | 32.9305136 | 30.7142857 | 2.21622788 | 0.55817789 | Chi-square |
| med_calcium_acetate | 195 | 31.9148936 | 32.9305136 | 30.7142857 | 2.21622788 | 0.55817789 | Chi-square |
| med_epinephrine | 195 | 31.9148936 | 32.9305136 | 30.7142857 | 2.21622788 | 0.55817789 | Chi-square |
| med_epinephrine_1:1000 | 195 | 31.9148936 | 32.9305136 | 30.7142857 | 2.21622788 | 0.55817789 | Chi-square |
| med_fentanyl_citrate | 195 | 31.9148936 | 32.9305136 | 30.7142857 | 2.21622788 | 0.55817789 | Chi-square |
| med_heparin | 195 | 31.9148936 | 32.9305136 | 30.7142857 | 2.21622788 | 0.55817789 | Chi-square |
| med_insulin | 195 | 31.9148936 | 32.9305136 | 30.7142857 | 2.21622788 | 0.55817789 | Chi-square |
| med_linezolid | 195 | 31.9148936 | 32.9305136 | 30.7142857 | 2.21622788 | 0.55817789 | Chi-square |
| med_mannitol | 195 | 31.9148936 | 32.9305136 | 30.7142857 | 2.21622788 | 0.55817789 | Chi-square |
| med_miconazole_2%_cream | 195 | 31.9148936 | 32.9305136 | 30.7142857 | 2.21622788 | 0.55817789 | Chi-square |
| med_midodrine | 195 | 31.9148936 | 32.9305136 | 30.7142857 | 2.21622788 | 0.55817789 | Chi-square |
| med_morphine_infusion_‚Äì_comfort_care_guidelines | 195 | 31.9148936 | 32.9305136 | 30.7142857 | 2.21622788 | 0.55817789 | Chi-square |
| med_nafcillin | 195 | 31.9148936 | 32.9305136 | 30.7142857 | 2.21622788 | 0.55817789 | Chi-square |
| med_nephrocaps | 195 | 31.9148936 | 32.9305136 | 30.7142857 | 2.21622788 | 0.55817789 | Chi-square |
| med_nitroprusside_sodium | 195 | 31.9148936 | 32.9305136 | 30.7142857 | 2.21622788 | 0.55817789 | Chi-square |
| med_norepinephrine | 195 | 31.9148936 | 32.9305136 | 30.7142857 | 2.21622788 | 0.55817789 | Chi-square |
| med_oxycodone_(immediate_release)_ | 195 | 31.9148936 | 32.9305136 | 30.7142857 | 2.21622788 | 0.55817789 | Chi-square |
| med_sarna_lotion | 195 | 31.9148936 | 32.9305136 | 30.7142857 | 2.21622788 | 0.55817789 | Chi-square |
| med_sodium_bicarbonate | 195 | 31.9148936 | 32.9305136 | 30.7142857 | 2.21622788 | 0.55817789 | Chi-square |
| med_tamsulosin | 195 | 31.9148936 | 32.9305136 | 30.7142857 | 2.21622788 | 0.55817789 | Chi-square |
| med_ursodiol | 195 | 31.9148936 | 32.9305136 | 30.7142857 | 2.21622788 | 0.55817789 | Chi-square |
| med_vasopressin | 195 | 31.9148936 | 32.9305136 | 30.7142857 | 2.21622788 | 0.55817789 | Chi-square |
| chart_first_spo2_desat_limit | 157 | 25.695581 | 34.7432024 | 15 | 19.7432024 | 2.62E-08 | Chi-square |
| output_first_spo2_desat_limit | 157 | 25.695581 | 34.7432024 | 15 | 19.7432024 | 2.62E-08 | Chi-square |
| chart_first_heart_rate_alarm_-_low | 139 | 22.7495908 | 32.0241692 | 11.7857143 | 20.2384549 | 2.75E-09 | Chi-square |
| chart_first_heart_rate_alarm_-_high | 138 | 22.5859247 | 31.7220544 | 11.7857143 | 19.9363401 | 4.30E-09 | Chi-square |
| lab_first_prothrombin_time | 131 | 21.4402619 | 25.9818731 | 16.0714286 | 9.91044454 | 0.00293883 | Chi-square |
| lab_first_platelet_count | 83 | 13.5842881 | 19.0332326 | 7.14285714 | 11.8903755 | 1.92E-05 | Chi-square |
| chart_first_hemoglobin | 77 | 12.6022913 | 17.2205438 | 7.14285714 | 10.0776867 | 0.0001841 | Chi-square |
| chart_first_glucose_(serum) | 69 | 11.2929624 | 17.8247734 | 3.57142857 | 14.2533448 | 2.92E-08 | Chi-square |
| chart_first_magnesium | 66 | 10.801964 | 16.6163142 | 3.92857143 | 12.6877428 | 4.80E-07 | Chi-square |
| chart_first_creatinine_(serum) | 62 | 10.1472995 | 16.0120846 | 3.21428571 | 12.7977989 | 1.79E-07 | Chi-square |
| lab_first_potassium_(serum) | 59 | 9.65630115 | 15.7099698 | 2.5 | 13.2099698 | 3.62E-08 | Chi-square |
| chart_first_respiratory_rate_(set) | 44 | 7.20130933 | 11.1782477 | 2.5 | 8.67824773 | 3.56E-05 | Chi-square |
| chart_first_respiratory_rate | 6 | 0.98199673 | 1.81268882 | 0 | 1.81268882 | 0.03374723 | Fisher |
| chart_first_heart_rate | 2 | 0.32733224 | 0.60422961 | 0 | 0.60422961 | 0.50266964 | Fisher |
| age | 0 | 0 | 0 | 0 | 0 |  | Failed |
| first_mGCS | 0 | 0 | 0 | 0 | 0 |  | Failed |
| gender | 0 | 0 | 0 | 0 | 0 |  | Failed |
| subject_id | 0 | 0 | 0 | 0 | 0 |  | Failed |
| underlying_cardiac_condition | 0 | 0 | 0 | 0 | 0 |  | Failed |

**eTable 17** – Complete feature list and missing variable report for MIMIC-IV dataset.

| **Variable** | **Missing, n** | **Missing, %** | **Missing in No-TTM Group, %** | **Missing in TTM Group, %** | **Absolute Difference, %** | **P Value** | **Statistical Test** |
| --- | --- | --- | --- | --- | --- | --- | --- |
| flo_first_r_amb_fcc_assess | 902 | 100 | 100 | 100 | 0 |  | Failed |
| flo_first_r_amb_fcc_total_time | 902 | 100 | 100 | 100 | 0 |  | Failed |
| flo_first_r_jhm_ad_masa_score | 902 | 100 | 100 | 100 | 0 |  | Failed |
| flo_first_r_jhm_ip_impella_motor_current_mean | 902 | 100 | 100 | 100 | 0 |  | Failed |
| flo_first_r_pca_patient_dose_mg | 902 | 100 | 100 | 100 | 0 |  | Failed |
| flo_first_r_rt_vt_vent_spontaneous | 902 | 100 | 100 | 100 | 0 |  | Failed |
| lab_first_blood_gases,pump_gas | 902 | 100 | 100 | 100 | 0 |  | Failed |
| lab_first_glucose,_pleural_fluid | 902 | 100 | 100 | 100 | 0 |  | Failed |
| lab_first_hematocrit | 902 | 100 | 100 | 100 | 0 |  | Failed |
| lab_first_hemoglobin | 902 | 100 | 100 | 100 | 0 |  | Failed |
| lab_first_hiv1_rna_realtime_pcr | 902 | 100 | 100 | 100 | 0 |  | Failed |
| lab_first_ph,_pleural_fluid | 902 | 100 | 100 | 100 | 0 |  | Failed |
| lab_first_poct_epoc_lactate | 902 | 100 | 100 | 100 | 0 |  | Failed |
| lab_first_prothrombin_time,superstat | 902 | 100 | 100 | 100 | 0 |  | Failed |
| lab_first_sun/creatinine_ratio | 902 | 100 | 100 | 100 | 0 |  | Failed |
| flo_first_r_cv_mac_av_hb_pa_value | 901 | 99.8891353 | 99.7867804 | 100 | 0.21321962 | 1 | Fisher |
| flo_first_r_cv_mac_av_hb_sa_value | 901 | 99.8891353 | 99.7867804 | 100 | 0.21321962 | 1 | Fisher |
| flo_first_r_head_3rd_degree_burn | 901 | 99.8891353 | 100 | 99.7690531 | 0.23094688 | 0.48004435 | Fisher |
| flo_first_r_ip_vent_flow_vcac | 901 | 99.8891353 | 99.7867804 | 100 | 0.21321962 | 1 | Fisher |
| flo_first_r_jh_blood_units_for_calculation | 901 | 99.8891353 | 100 | 99.7690531 | 0.23094688 | 0.48004435 | Fisher |
| flo_first_r_jhm_an_perf_perfusion_flow | 901 | 99.8891353 | 99.7867804 | 100 | 0.21321962 | 1 | Fisher |
| flo_first_r_jhm_ip_cerebral_oximetry_right | 901 | 99.8891353 | 99.7867804 | 100 | 0.21321962 | 1 | Fisher |
| flo_first_r_jhm_ip_crrt_circuit_volume | 901 | 99.8891353 | 100 | 99.7690531 | 0.23094688 | 0.48004435 | Fisher |
| flo_first_r_jhm_ip_ecmo_pump_speed | 901 | 99.8891353 | 100 | 99.7690531 | 0.23094688 | 0.48004435 | Fisher |
| flo_first_r_jhm_ip_icu_cards_scale_score | 901 | 99.8891353 | 99.7867804 | 100 | 0.21321962 | 1 | Fisher |
| flo_first_r_jhm_perf_cerebral_perfusion_temp | 901 | 99.8891353 | 99.7867804 | 100 | 0.21321962 | 1 | Fisher |
| flo_first_r_l_thigh_circumference_(cm) | 901 | 99.8891353 | 100 | 99.7690531 | 0.23094688 | 0.48004435 | Fisher |
| flo_first_r_left_buttock_2nd_degree_burn | 901 | 99.8891353 | 99.7867804 | 100 | 0.21321962 | 1 | Fisher |
| flo_first_r_neck_2nd_degree_burn | 901 | 99.8891353 | 99.7867804 | 100 | 0.21321962 | 1 | Fisher |
| flo_first_r_perf_gas_flow | 901 | 99.8891353 | 99.7867804 | 100 | 0.21321962 | 1 | Fisher |
| flo_first_r_perf_venous_temp | 901 | 99.8891353 | 99.7867804 | 100 | 0.21321962 | 1 | Fisher |
| flo_first_r_pt_treatment_time | 901 | 99.8891353 | 99.7867804 | 100 | 0.21321962 | 1 | Fisher |
| flo_first_r_right_buttock_3rd_degree_burn | 901 | 99.8891353 | 100 | 99.7690531 | 0.23094688 | 0.48004435 | Fisher |
| lab_first_calcium,_serum | 901 | 99.8891353 | 99.7867804 | 100 | 0.21321962 | 1 | Fisher |
| lab_first_ige,_serum | 901 | 99.8891353 | 99.7867804 | 100 | 0.21321962 | 1 | Fisher |
| lab_first_platelet_function_test | 901 | 99.8891353 | 99.7867804 | 100 | 0.21321962 | 1 | Fisher |
| lab_first_protein,_peritoneal_fluid | 901 | 99.8891353 | 100 | 99.7690531 | 0.23094688 | 0.48004435 | Fisher |
| lab_first_sodium | 901 | 99.8891353 | 100 | 99.7690531 | 0.23094688 | 0.48004435 | Fisher |
| lab_first_stroke_aptt | 901 | 99.8891353 | 99.7867804 | 100 | 0.21321962 | 1 | Fisher |
| lab_first_stroke_troponin | 901 | 99.8891353 | 99.7867804 | 100 | 0.21321962 | 1 | Fisher |
| lab_first_thromboelastograph,_rapid-jhh_only | 901 | 99.8891353 | 99.7867804 | 100 | 0.21321962 | 1 | Fisher |
| lab_first_total_protein_(body_fluid) | 901 | 99.8891353 | 100 | 99.7690531 | 0.23094688 | 0.48004435 | Fisher |
| lab_first_tryptase | 901 | 99.8891353 | 99.7867804 | 100 | 0.21321962 | 1 | Fisher |
| flo_first_*old_r_jhm_ip_qtc_interval | 900 | 99.7782705 | 99.7867804 | 99.7690531 | 0.01772727 | 1 | Fisher |
| flo_first_r_an_spo2 | 900 | 99.7782705 | 99.7867804 | 99.7690531 | 0.01772727 | 1 | Fisher |
| flo_first_r_cpn_glasgow_coma_scale_score_2 | 900 | 99.7782705 | 100 | 99.5381062 | 0.46189376 | 0.23016555 | Fisher |
| flo_first_r_cv_mac_blood_temp | 900 | 99.7782705 | 99.5735608 | 100 | 0.42643923 | 0.5002424 | Fisher |
| flo_first_r_dobutamine_volume | 900 | 99.7782705 | 99.7867804 | 99.7690531 | 0.01772727 | 1 | Fisher |
| lab_first_renal_function_panel | 900 | 99.7782705 | 99.5735608 | 100 | 0.42643923 | 0.5002424 | Fisher |
| flo_first_*old_r_ed_sedation_narr_qt_interval | 899 | 99.6674058 | 100 | 99.3071594 | 0.69284065 | 0.11022372 | Fisher |
| flo_first_r_blood_glucose_meter | 899 | 99.6674058 | 99.7867804 | 99.5381062 | 0.24867415 | 0.61018907 | Fisher |
| flo_first_r_jhm_ip_impella_left_motor_current_mean | 899 | 99.6674058 | 99.3603412 | 100 | 0.63965885 | 0.2503636 | Fisher |
| flo_first_r_lorazepam_volume | 899 | 99.6674058 | 99.7867804 | 99.5381062 | 0.24867415 | 0.61018907 | Fisher |
| lab_first_b-type_natriuretic_peptide | 899 | 99.6674058 | 99.7867804 | 99.5381062 | 0.24867415 | 0.61018907 | Fisher |
| lab_first_vitamin_d_(25-hydroxy)_total | 899 | 99.6674058 | 99.7867804 | 99.5381062 | 0.24867415 | 0.61018907 | Fisher |
| flo_first_r_cerebral_perfusion_pressure | 898 | 99.556541 | 99.5735608 | 99.5381062 | 0.03545453 | 1 | Fisher |
| flo_first_r_ip_vent_vt_low | 898 | 99.556541 | 99.5735608 | 99.5381062 | 0.03545453 | 1 | Fisher |
| flo_first_r_jhm_ip_temp_goal_2 | 898 | 99.556541 | 99.7867804 | 99.3071594 | 0.47962103 | 0.35537385 | Fisher |
| flo_first_r_pulmonary_capillary_wedge_pressure | 898 | 99.556541 | 99.1471215 | 100 | 0.85287846 | 0.12536305 | Fisher |
| flo_first_r_pulmonary_vasclar_resistance_calc | 898 | 99.556541 | 99.1471215 | 100 | 0.85287846 | 0.12536305 | Fisher |
| lab_first_haptoglobin | 898 | 99.556541 | 99.3603412 | 99.7690531 | 0.40871197 | 0.62536525 | Fisher |
| lab_first_platelet_(citrate)_count | 898 | 99.556541 | 99.5735608 | 99.5381062 | 0.03545453 | 1 | Fisher |
| flo_first_r_morse_fall_risk_score | 897 | 99.4456763 | 100 | 98.8452656 | 1.15473441 | 0.02518632 | Fisher |
| flo_first_r_systemic_vascular_resistance_calc | 897 | 99.4456763 | 98.9339019 | 100 | 1.06609808 | 0.06280163 | Fisher |
| flo_first_r_total_burn_area | 897 | 99.4456763 | 99.1471215 | 99.7690531 | 0.62193158 | 0.37560875 | Fisher |
| lab_first_potassium | 896 | 99.3348115 | 99.3603412 | 99.3071594 | 0.0531818 | 1 | Fisher |
| lab_first_hepatic_function_panel | 894 | 99.113082 | 99.3603412 | 98.8452656 | 0.51507556 | 0.49100975 | Fisher |
| lab_first_platelet_count | 894 | 99.113082 | 100 | 98.1524249 | 1.84757506 | 0.00272598 | Fisher |
| lab_first_thromboelastograph_clotting_profile | 894 | 99.113082 | 98.9339019 | 99.3071594 | 0.37325743 | 0.72709999 | Fisher |
| flo_first_pulse_ox_heart_rate | 893 | 99.0022173 | 98.5074627 | 99.5381062 | 1.03064355 | 0.18064626 | Fisher |
| flo_first_emesis | 892 | 98.8913525 | 99.5735608 | 98.1524249 | 1.42113583 | 0.05550322 | Fisher |
| flo_first_r_an_agents_n2o_flow | 892 | 98.8913525 | 98.2942431 | 99.5381062 | 1.24386317 | 0.11022714 | Fisher |
| lab_first_c-reactive_protein | 892 | 98.8913525 | 98.5074627 | 99.3071594 | 0.79969667 | 0.34495753 | Fisher |
| lab_first_sedimentation_rate | 892 | 98.8913525 | 98.9339019 | 98.8452656 | 0.08863633 | 1 | Fisher |
| flo_first_r_paco2 | 890 | 98.6696231 | 99.1471215 | 98.1524249 | 0.99469659 | 0.19268311 | Chi-square |
| flo_first_r_resp_ph | 890 | 98.6696231 | 99.1471215 | 98.1524249 | 0.99469659 | 0.19268311 | Chi-square |
| flo_first_r_resp_vt_set/target_ml | 890 | 98.6696231 | 98.9339019 | 98.3833718 | 0.55053009 | 0.47091584 | Chi-square |
| flo_first_r_sao2 | 890 | 98.6696231 | 99.1471215 | 98.1524249 | 0.99469659 | 0.19268311 | Chi-square |
| lab_first_glucose,_blood-_poc | 890 | 98.6696231 | 99.1471215 | 98.1524249 | 0.99469659 | 0.19268311 | Chi-square |
| lab_first_coagulation_screen | 889 | 98.5587583 | 99.7867804 | 97.2286374 | 2.55814297 | 0.00127926 | Chi-square |
| flo_first_r_pao2 | 888 | 98.4478936 | 99.1471215 | 97.6905312 | 1.45659036 | 0.07704844 | Chi-square |
| flo_first_r_jhm_ip_qtc_ainterval_(sec) | 887 | 98.3370288 | 99.3603412 | 97.2286374 | 2.13170374 | 0.01237639 | Chi-square |
| flo_first_r_ad_left_pupil_npi | 885 | 98.1152993 | 97.228145 | 99.0762125 | 1.84806748 | 0.04143216 | Chi-square |
| flo_first_r_ad_right_pupil_npi | 885 | 98.1152993 | 97.228145 | 99.0762125 | 1.84806748 | 0.04143216 | Chi-square |
| flo_first_anesthesia_temperature | 883 | 97.8935698 | 96.8017058 | 99.0762125 | 2.27450671 | 0.01747105 | Chi-square |
| flo_first_r_cardiac_index | 883 | 97.8935698 | 98.5074627 | 97.2286374 | 1.27882527 | 0.18146759 | Chi-square |
| lab_first_ferritin | 881 | 97.6718404 | 97.8678038 | 97.4595843 | 0.40821954 | 0.68460259 | Chi-square |
| lab_first_ck_mb_reflex | 873 | 96.7849224 | 97.8678038 | 95.6120092 | 2.2557946 | 0.05501113 | Chi-square |
| flo_first_r_ip_blood_administration_volume | 872 | 96.6740576 | 96.5884861 | 96.7667436 | 0.17825751 | 0.88142461 | Chi-square |
| lab_first_poct_istat_lactate | 872 | 96.6740576 | 97.4413646 | 95.8429561 | 1.59840849 | 0.18105069 | Chi-square |
| flo_first_r_an_fio2 | 867 | 96.1197339 | 94.8827292 | 97.4595843 | 2.57685508 | 0.0452768 | Chi-square |
| lab_first_urinalysis_with_reflex_culture | 865 | 95.8980044 | 95.7356077 | 96.073903 | 0.33829533 | 0.79800511 | Chi-square |
| flo_first_r_jhh_post-treatment_rr | 855 | 94.789357 | 96.1620469 | 93.3025404 | 2.85950649 | 0.0535323 | Chi-square |
| flo_first_r_vasopressin_volume | 849 | 94.1241685 | 95.9488273 | 92.147806 | 3.80102129 | 0.01530123 | Chi-square |
| flo_first_r_ip_vent_vt_high | 839 | 93.0155211 | 94.2430704 | 91.6859122 | 2.55715812 | 0.13223191 | Chi-square |
| lab_first_d-dimer,_quantitative | 839 | 93.0155211 | 94.2430704 | 91.6859122 | 2.55715812 | 0.13223191 | Chi-square |
| flo_first_r_cv_mac_spo2 | 830 | 92.0177384 | 91.8976546 | 92.147806 | 0.25015142 | 0.88984992 | Chi-square |
| flo_first_r_amiodarone_volume | 818 | 90.6873614 | 90.6183369 | 90.7621247 | 0.14378782 | 0.94081922 | Chi-square |
| flo_first_r_jhm_ip_sofa_resp_score | 805 | 89.2461197 | 89.978678 | 88.4526559 | 1.52602215 | 0.45983856 | Chi-square |
| flo_first_intravenous_intake | 794 | 88.0266075 | 89.5522388 | 86.3741339 | 3.17810486 | 0.14187138 | Chi-square |
| lab_first_hemoglobin_a1c | 791 | 87.6940133 | 85.9275053 | 89.6073903 | 3.67988497 | 0.09280018 | Chi-square |
| flo_first_r_jhm_ip_temp_goal | 751 | 83.2594235 | 88.2729211 | 77.8290993 | 10.4438218 | 2.70E-05 | Chi-square |
| lab_first_creatine_kinase_(ck) | 745 | 82.594235 | 85.5010661 | 79.4457275 | 6.05533862 | 0.0165608 | Chi-square |
| lab_first_sodium,whole_blood | 738 | 81.8181818 | 79.3176972 | 84.5265589 | 5.20886166 | 0.04272325 | Chi-square |
| lab_first_lipase | 735 | 81.4855876 | 84.8614072 | 77.8290993 | 7.03230794 | 0.00659514 | Chi-square |
| lab_first_pro-b_natriuretic_peptide | 722 | 80.0443459 | 79.1044776 | 81.0623557 | 1.95787805 | 0.46231158 | Chi-square |
| flo_first_r_ecg_lead3_st | 704 | 78.0487805 | 75.0533049 | 81.2933025 | 6.23999764 | 0.0236953 | Chi-square |
| flo_first_r_ecg_lead1_st | 703 | 77.9379157 | 74.8400853 | 81.2933025 | 6.45321725 | 0.01953813 | Chi-square |
| flo_first_r_ecg_lead2_st | 703 | 77.9379157 | 75.0533049 | 81.0623557 | 6.00905075 | 0.02967666 | Chi-square |
| lab_first_calcium,_ionized,_whole_blood | 667 | 73.9467849 | 73.9872068 | 73.9030023 | 0.08420451 | 0.97703575 | Chi-square |
| flo_first_*old_r_pupil_size_(mm)_left | 616 | 68.2926829 | 70.3624733 | 66.0508083 | 4.31166503 | 0.16444095 | Chi-square |
| flo_first_r_jhm_passes | 614 | 68.0709534 | 69.9360341 | 66.0508083 | 3.8852258 | 0.21113191 | Chi-square |
| lab_first_complete_blood_count_(cbc)_+_auto_diff | 609 | 67.5166297 | 65.8848614 | 69.2840647 | 3.39920326 | 0.27610917 | Chi-square |
| lab_first_troponin | 578 | 64.0798226 | 64.8187633 | 63.2794457 | 1.5393176 | 0.63021699 | Chi-square |
| lab_first_magnesium | 527 | 58.4257206 | 61.1940299 | 55.4272517 | 5.76677812 | 0.07914267 | Chi-square |
| flo_first_r_jhm_ip_sofa_liver_score | 493 | 54.6563193 | 55.4371002 | 53.8106236 | 1.62647666 | 0.62397397 | Chi-square |
| flo_first_r_jhm_ip_sofa_cv_score | 485 | 53.7694013 | 55.8635394 | 51.5011547 | 4.36238471 | 0.18922983 | Chi-square |
| flo_first_r_jhm_ip_sofa_renal_score | 481 | 53.3259424 | 54.3710021 | 52.1939954 | 2.17700675 | 0.51262453 | Chi-square |
| flo_first_r_jhm_ip_sofa_coagulation_score | 478 | 52.9933481 | 54.3710021 | 51.5011547 | 2.8698474 | 0.38826336 | Chi-square |
| flo_first_r_sofa_score | 462 | 51.2195122 | 52.238806 | 50.1154734 | 2.12333253 | 0.52387172 | Chi-square |
| lab_first_prothrombin_time_+_inr | 448 | 49.6674058 | 50.533049 | 48.7297921 | 1.80325689 | 0.58839849 | Chi-square |
| flo_first_haz_r_bmi_osa_screen | 412 | 45.6762749 | 47.3347548 | 43.8799076 | 3.45484718 | 0.29802562 | Chi-square |
| flo_first_r_bmi | 412 | 45.6762749 | 47.3347548 | 43.8799076 | 3.45484718 | 0.29802562 | Chi-square |
| flo_first_height | 333 | 36.9179601 | 39.8720682 | 33.7182448 | 6.15382343 | 0.05569988 | Chi-square |
| flo_first_r_jhm_ip_rt_conv._vent._low_respiratory_rate | 310 | 34.368071 | 35.8208955 | 32.7944573 | 3.02643825 | 0.33899899 | Chi-square |
| flo_first_r_jhm_ip_rt_conv._vent._high_respiratory_rate | 252 | 27.9379157 | 28.7846482 | 27.0207852 | 1.76386297 | 0.55529127 | Chi-square |
| flo_first_r_fio2 | 209 | 23.1707317 | 24.3070362 | 21.9399538 | 2.36708244 | 0.39990171 | Chi-square |
| flo_first_r_jhm_ip_weight_kg | 194 | 21.5077605 | 21.5351812 | 21.47806 | 0.05712119 | 0.98335737 | Chi-square |
| flo_first_r_skin_braden_scale_score | 167 | 18.5144124 | 19.8294243 | 17.0900693 | 2.73935502 | 0.28995061 | Chi-square |
| flo_first_r_cpn_glasgow_coma_scale_score | 127 | 14.0798226 | 15.3518124 | 12.7020785 | 2.64973384 | 0.25299822 | Chi-square |
| flo_first_weight/scale | 116 | 12.8603104 | 15.565032 | 9.93071594 | 5.63431605 | 0.01155583 | Chi-square |
| flo_first_r_apache_temperature | 46 | 5.09977827 | 4.47761194 | 5.77367206 | 1.29606012 | 0.37670641 | Chi-square |
| flo_first_pulse | 4 | 0.44345898 | 0.85287846 | 0 | 0.85287846 | 0.12536305 | Fisher |
| VF | 0 | 0 | 0 | 0 | 0 |  | Failed |
| age | 0 | 0 | 0 | 0 | 0 |  | Failed |
| asystole | 0 | 0 | 0 | 0 | 0 |  | Failed |
| cardiac arrest with successful resuscitation | 0 | 0 | 0 | 0 | 0 |  | Failed |
| cardiopulmonary arrest | 0 | 0 | 0 | 0 | 0 |  | Failed |
| cardiopulmonary arrest w/ resuscitation | 0 | 0 | 0 | 0 | 0 |  | Failed |
| death due to cardiac arrest | 0 | 0 | 0 | 0 | 0 |  | Failed |
| ed_visit_yn | 0 | 0 | 0 | 0 | 0 |  | Failed |
| flo_first_bp_diastolic | 0 | 0 | 0 | 0 | 0 |  | Failed |
| flo_jhm_ip_cuff_pressure_string_Tachycardia | 0 | 0 | 0 | 0 | 0 |  | Failed |
| flo_jhm_ip_cust_formula_fall_risk_0-No fall history w/in 6 months prior to admission | 0 | 0 | 0 | 0 | 0 |  | Failed |
| flo_jhm_r_ed_electronic_triage_supplement_Low Risk - Paralysis/Completely Immobilized | 0 | 0 | 0 | 0 | 0 |  | Failed |
| flo_r_an_red_screen_monitor_Asytole - Brady at 30- Asystole - PEA - Asystole | 0 | 0 | 0 | 0 | 0 |  | Failed |
| flo_r_an_red_screen_monitor_Atrial fibrillation | 0 | 0 | 0 | 0 | 0 |  | Failed |
| flo_r_an_red_screen_monitor_Calcium and bicarb given IV | 0 | 0 | 0 | 0 | 0 |  | Failed |
| flo_r_an_red_screen_monitor_Mary Carter | 0 | 0 | 0 | 0 | 0 |  | Failed |
| flo_r_cam-icu_overall_ M16 | 0 | 0 | 0 | 0 | 0 |  | Failed |
| flo_r_cam-icu_overall_BC19 | 0 | 0 | 0 | 0 | 0 |  | Failed |
| flo_r_cam-icu_overall_Positional | 0 | 0 | 0 | 0 | 0 |  | Failed |
| flo_r_ed_care_handoff_rn_14 | 0 | 0 | 0 | 0 | 0 |  | Failed |
| flo_r_ed_ecg_proc_md_name_0-No mobility issues | 0 | 0 | 0 | 0 | 0 |  | Failed |
| flo_r_ed_ecg_proc_md_name_Continuously | 0 | 0 | 0 | 0 | 0 |  | Failed |
| flo_r_ed_ecg_proc_md_name_Flushed | 0 | 0 | 0 | 0 | 0 |  | Failed |
| flo_r_ed_ecg_proc_md_name_Gabor Kelen | 0 | 0 | 0 | 0 | 0 |  | Failed |
| flo_r_ed_ecg_proc_md_name_Tammy Waldman | 0 | 0 | 0 | 0 | 0 |  | Failed |
| flo_r_ed_ecg_proc_md_name_Tan | 0 | 0 | 0 | 0 | 0 |  | Failed |
| flo_r_ed_ecg_proc_md_name_Tara RN | 0 | 0 | 0 | 0 | 0 |  | Failed |
| flo_r_ed_jhh_plan_of_care_notes_Age > 65 | 0 | 0 | 0 | 0 | 0 |  | Failed |
| flo_r_ed_jhh_plan_of_care_notes_Hypertension | 0 | 0 | 0 | 0 | 0 |  | Failed |
| flo_r_ed_pre-hospital_ekg_no cardiac activity on bedside echo | 0 | 0 | 0 | 0 | 0 |  | Failed |
| flo_r_ed_vitals_assessment_timer_Critical | 0 | 0 | 0 | 0 | 0 |  | Failed |
| flo_r_ed_vitals_assessment_timer_Heart block | 0 | 0 | 0 | 0 | 0 |  | Failed |
| flo_r_ed_vitals_assessment_timer_Mental Status Change | 0 | 0 | 0 | 0 | 0 |  | Failed |
| flo_r_ed_vitals_assessment_timer_Original surgical dressing intact | 0 | 0 | 0 | 0 | 0 |  | Failed |
| flo_r_ed_vitals_assessment_timer_Unable to return demo and/or verbal instructions | 0 | 0 | 0 | 0 | 0 |  | Failed |
| flo_r_ed_vitals_assessment_timer_bicarb givent | 0 | 0 | 0 | 0 | 0 |  | Failed |
| flo_r_ed_vitals_assessment_timer_shock delivered contiuned cpr | 0 | 0 | 0 | 0 | 0 |  | Failed |
| flo_r_ip_fall_risk_category_Pos | 0 | 0 | 0 | 0 | 0 |  | Failed |
| flo_r_ip_fall_risk_category_Voice | 0 | 0 | 0 | 0 | 0 |  | Failed |
| flo_r_ip_rt_jhm_conv._vent_high_tv_plan for Ca, mag, bicarb, amio | 0 | 0 | 0 | 0 | 0 |  | Failed |
| flo_r_jhh_ed_dispo_recommendation_text_Pt with history of seizure and on suboxaone | 0 | 0 | 0 | 0 | 0 |  | Failed |
| flo_r_jhh_ed_dispo_recommendation_text_cardiac surgery at bedside | 0 | 0 | 0 | 0 | 0 |  | Failed |
| flo_r_jhh_ed_dispo_supplemental_info_no cardiac motion on bedside echo | 0 | 0 | 0 | 0 | 0 |  | Failed |
| flo_r_jhh_ip_rt_low_etco2_alarm_Verbal | 0 | 0 | 0 | 0 | 0 |  | Failed |
| flo_r_jhm_ed_ambulance_run_49 | 0 | 0 | 0 | 0 | 0 |  | Failed |
| flo_r_jhm_ed_ambulance_run_Right brachial | 0 | 0 | 0 | 0 | 0 |  | Failed |
| flo_r_jhm_ed_dispo_acute_risk_cardiac arrest | 0 | 0 | 0 | 0 | 0 |  | Failed |
| flo_r_jhm_ed_dispo_crit_risk_Pt arrived. Unwitnessed arrest, pt paraplegic, recent RLE surgery, heroin abuse | 0 | 0 | 0 | 0 | 0 |  | Failed |
| flo_r_jhm_ed_fall_risk_custom_formula_Patient has stopped bleeding @ this time. Plan to have 6.0 cuffed tube placed. Neuro IR paged | 0 | 0 | 0 | 0 | 0 |  | Failed |
| flo_r_jhm_ip_rt_i:e_2_Body tag and bag tag match patient ID band | 0 | 0 | 0 | 0 | 0 |  | Failed |
| flo_r_jhm_ip_rt_mech_vent_id_#_provider doing bedsaide ultrasound | 0 | 0 | 0 | 0 | 0 |  | Failed |
| flo_r_jhm_ip_rt_observed_i:e_DC med31 - 229102 | 0 | 0 | 0 | 0 | 0 |  | Failed |
| flo_r_jhm_ip_rt_observed_i:e_central line and arterial line to be placed per Dr. Risko. | 0 | 0 | 0 | 0 | 0 |  | Failed |
| flo_r_jhm_rcp_time_Passive | 0 | 0 | 0 | 0 | 0 |  | Failed |
| flo_r_jhm_trews_session_end_dtm_Zoll pads on pt. | 0 | 0 | 0 | 0 | 0 |  | Failed |
| flo_r_jhm_trews_session_manual_override_History of DM | 0 | 0 | 0 | 0 | 0 |  | Failed |
| flo_r_jhm_trews_version_1# | 0 | 0 | 0 | 0 | 0 |  | Failed |
| flo_r_jhm_trews_version_105 | 0 | 0 | 0 | 0 | 0 |  | Failed |
| flo_r_jhm_trews_version_11 | 0 | 0 | 0 | 0 | 0 |  | Failed |
| flo_r_jhm_trews_version_1:1.3 | 0 | 0 | 0 | 0 | 0 |  | Failed |
| flo_r_jhm_trews_version_Incoming cardiac arrest | 0 | 0 | 0 | 0 | 0 |  | Failed |
| flo_r_jhm_trews_version_Irregular | 0 | 0 | 0 | 0 | 0 |  | Failed |
| flo_r_non-violent_describe_behaviors_Yes | 0 | 0 | 0 | 0 | 0 |  | Failed |
| flo_r_provider_name_ Adult Neurosurgery - Low Risk | 0 | 0 | 0 | 0 | 0 |  | Failed |
| med_amiodarone_150_mg/100_ml | 0 | 0 | 0 | 0 | 0 |  | Failed |
| med_amiodarone_360_mg/200_ml | 0 | 0 | 0 | 0 | 0 |  | Failed |
| med_chlorhexidine_gluconate_0.12% | 0 | 0 | 0 | 0 | 0 |  | Failed |
| med_dapsone_100_mg_tablet | 0 | 0 | 0 | 0 | 0 |  | Failed |
| med_epinephrine_0.1_mg/ml_injection_syringe | 0 | 0 | 0 | 0 | 0 |  | Failed |
| med_epinephrine_1_mg/ml | 0 | 0 | 0 | 0 | 0 |  | Failed |
| med_epinephrine_hcl_2_mg/100_ml | 0 | 0 | 0 | 0 | 0 |  | Failed |
| med_epinephrine_infusion_4_mcg/ml_in_250_ml_d5w | 0 | 0 | 0 | 0 | 0 |  | Failed |
| med_famotidine | 0 | 0 | 0 | 0 | 0 |  | Failed |
| med_heparin | 0 | 0 | 0 | 0 | 0 |  | Failed |
| med_lidocaine | 0 | 0 | 0 | 0 | 0 |  | Failed |
| med_lorazepam_2_mg/ml_injection_syringe | 0 | 0 | 0 | 0 | 0 |  | Failed |
| med_morphine_10_mg/ml_injection_syringe | 0 | 0 | 0 | 0 | 0 |  | Failed |
| med_morphine_10_mg/ml_soln_erx_5168 | 0 | 0 | 0 | 0 | 0 |  | Failed |
| med_norepinephrine_infusion_16_mg_/_116_ml_ns | 0 | 0 | 0 | 0 | 0 |  | Failed |
| med_norepinephrine_infusion_8_mg_/_250_ml_ns | 0 | 0 | 0 | 0 | 0 |  | Failed |
| med_ondansetron_hcl | 0 | 0 | 0 | 0 | 0 |  | Failed |
| med_sodium_bicarbonate_8.4_% | 0 | 0 | 0 | 0 | 0 |  | Failed |
| med_sodium_bicarbonate_infusion | 0 | 0 | 0 | 0 | 0 |  | Failed |
| med_sodium_chloride_bolus | 0 | 0 | 0 | 0 | 0 |  | Failed |
| pea | 0 | 0 | 0 | 0 | 0 |  | Failed |
| traumatic cardiac arrest | 0 | 0 | 0 | 0 | 0 |  | Failed |

**eTable 18** – Complete feature list and missing variable report for PMAP dataset.

| **Variable** | **Missing, n** | **Missing, %** | **Missing in No-TTM Group, %** | **Missing in TTM Group, %** | **Absolute Difference, %** | **P Value** | **Statistical Test** |
| --- | --- | --- | --- | --- | --- | --- | --- |
| EI_TRANSFUS | 564 | 97.0740103 | 97.3063973 | 96.8309859 | 0.47541139 | 0.7339437 | Chi-square |
| EI_INTRACER | 564 | 97.0740103 | 97.3063973 | 96.8309859 | 0.47541139 | 0.7339437 | Chi-square |
| EI_CHIR | 564 | 97.0740103 | 97.3063973 | 96.8309859 | 0.47541139 | 0.7339437 | Chi-square |
| V0_ANGIO_YES | 538 | 92.5989673 | 92.5925926 | 92.6056338 | 0.01304121 | 0.99521088 | Chi-square |
| J0_BICARB_DOS | 533 | 91.7383821 | 91.9191919 | 91.5492958 | 0.36989614 | 0.87139218 | Chi-square |
| BIO_TROPO2 | 521 | 89.6729776 | 88.8888889 | 90.4929577 | 1.60406886 | 0.52535188 | Chi-square |
| BIO_TROPO_CGT | 520 | 89.5008606 | 89.2255892 | 89.7887324 | 0.56314317 | 0.82482065 | Chi-square |
| V0_CHARLSON18 | 507 | 87.2633391 | 86.5319865 | 88.028169 | 1.49618248 | 0.58868535 | Chi-square |
| EI_ECHO | 466 | 80.2065404 | 78.1144781 | 82.3943662 | 4.27988808 | 0.19558132 | Chi-square |
| EI_DIURETIQ | 465 | 80.0344234 | 78.1144781 | 82.0422535 | 3.92777541 | 0.23645091 | Chi-square |
| EI_ANTIEPILEPTIQ | 447 | 76.9363167 | 73.4006734 | 80.6338028 | 7.23312942 | 0.03855277 | Chi-square |
| EI_ARYTHMI | 421 | 72.4612737 | 69.6969697 | 75.3521127 | 5.65514298 | 0.12717313 | Chi-square |
| EI_HEMOSEVER | 420 | 72.2891566 | 69.6969697 | 75 | 5.3030303 | 0.15340187 | Chi-square |
| EI_EXTRARENAL | 420 | 72.2891566 | 69.6969697 | 75 | 5.3030303 | 0.15340187 | Chi-square |
| EI_OAP | 420 | 72.2891566 | 69.6969697 | 75 | 5.3030303 | 0.15340187 | Chi-square |
| EI_CONVULS | 420 | 72.2891566 | 69.6969697 | 75 | 5.3030303 | 0.15340187 | Chi-square |
| J0_IRC | 377 | 64.8881239 | 63.6363636 | 66.1971831 | 2.56081946 | 0.51800365 | Chi-square |
| J0_HYPERCAP | 377 | 64.8881239 | 63.6363636 | 66.1971831 | 2.56081946 | 0.51800365 | Chi-square |
| J0_O2 | 377 | 64.8881239 | 63.6363636 | 66.1971831 | 2.56081946 | 0.51800365 | Chi-square |
| J0_TABAC | 377 | 64.8881239 | 63.6363636 | 66.1971831 | 2.56081946 | 0.51800365 | Chi-square |
| BIO_DDIMERE | 372 | 64.0275387 | 62.962963 | 65.1408451 | 2.17788211 | 0.58452764 | Chi-square |
| J0_DSA_P | 371 | 63.8554217 | 64.3097643 | 63.3802817 | 0.92948262 | 0.81567214 | Chi-square |
| BIO_LIPAS | 344 | 59.2082616 | 57.2390572 | 61.2676056 | 4.02854839 | 0.3233045 | Chi-square |
| BIO_MAGNE | 315 | 54.2168675 | 54.8821549 | 53.5211268 | 1.36102812 | 0.74204082 | Chi-square |
| J0_OCULAIRE | 297 | 51.1187608 | 50.1683502 | 52.1126761 | 1.94432589 | 0.63931327 | Chi-square |
| J0_VERBALE | 297 | 51.1187608 | 50.1683502 | 52.1126761 | 1.94432589 | 0.63931327 | Chi-square |
| J0_MOTRICE | 297 | 51.1187608 | 50.1683502 | 52.1126761 | 1.94432589 | 0.63931327 | Chi-square |
| J0_GLASGOW | 296 | 50.9466437 | 49.8316498 | 52.1126761 | 2.28102622 | 0.58247157 | Chi-square |
| J0_NYHA | 238 | 40.9638554 | 39.3939394 | 42.6056338 | 3.21169441 | 0.43133554 | Chi-square |
| J0_MYOCARD | 238 | 40.9638554 | 39.3939394 | 42.6056338 | 3.21169441 | 0.43133554 | Chi-square |
| J0_ARTERIO | 238 | 40.9638554 | 39.3939394 | 42.6056338 | 3.21169441 | 0.43133554 | Chi-square |
| J0_HTA | 238 | 40.9638554 | 39.3939394 | 42.6056338 | 3.21169441 | 0.43133554 | Chi-square |
| J0_NORA | 236 | 40.6196213 | 38.3838384 | 42.9577465 | 4.5739081 | 0.26180259 | Chi-square |
| J0_ADRE2 | 236 | 40.6196213 | 38.3838384 | 42.9577465 | 4.5739081 | 0.26180259 | Chi-square |
| J0_DOBU | 236 | 40.6196213 | 38.3838384 | 42.9577465 | 4.5739081 | 0.26180259 | Chi-square |
| J0_DOPA | 236 | 40.6196213 | 38.3838384 | 42.9577465 | 4.5739081 | 0.26180259 | Chi-square |
| J0_REFLEXVEST | 231 | 39.7590361 | 38.047138 | 41.5492958 | 3.50215773 | 0.38856313 | Chi-square |
| ECG_SALV_SUPRA | 228 | 39.242685 | 39.0572391 | 39.4366197 | 0.37938066 | 0.92541492 | Chi-square |
| ECG_SUS_ST | 227 | 39.070568 | 39.0572391 | 39.084507 | 0.02726799 | 0.9946272 | Chi-square |
| ECG_SOUS_ST | 227 | 39.070568 | 39.0572391 | 39.084507 | 0.02726799 | 0.9946272 | Chi-square |
| ECG_BAVI | 227 | 39.070568 | 39.0572391 | 39.084507 | 0.02726799 | 0.9946272 | Chi-square |
| ECG_BAVII | 227 | 39.070568 | 39.0572391 | 39.084507 | 0.02726799 | 0.9946272 | Chi-square |
| ECG_BAVIII | 227 | 39.070568 | 39.0572391 | 39.084507 | 0.02726799 | 0.9946272 | Chi-square |
| ECG_BBG | 227 | 39.070568 | 39.0572391 | 39.084507 | 0.02726799 | 0.9946272 | Chi-square |
| ECG_BBD | 227 | 39.070568 | 39.0572391 | 39.084507 | 0.02726799 | 0.9946272 | Chi-square |
| ECG_TACHICARD | 227 | 39.070568 | 39.0572391 | 39.084507 | 0.02726799 | 0.9946272 | Chi-square |
| ECG_FIBRIL | 227 | 39.070568 | 39.0572391 | 39.084507 | 0.02726799 | 0.9946272 | Chi-square |
| ECG_SALV_VENT | 227 | 39.070568 | 39.0572391 | 39.084507 | 0.02726799 | 0.9946272 | Chi-square |
| ECG_FLUTER | 227 | 39.070568 | 39.0572391 | 39.084507 | 0.02726799 | 0.9946272 | Chi-square |
| J0_REFLEXCARD | 182 | 31.3253012 | 31.3131313 | 31.3380282 | 0.02489686 | 0.99483958 | Chi-square |
| J0_DSA | 174 | 29.9483649 | 28.956229 | 30.9859155 | 2.02968654 | 0.59339143 | Chi-square |
| J0_CARDIO | 147 | 25.3012048 | 22.8956229 | 27.8169014 | 4.92127851 | 0.1725819 | Chi-square |
| J0_POUMON | 147 | 25.3012048 | 22.8956229 | 27.8169014 | 4.92127851 | 0.1725819 | Chi-square |
| J0_REFLEXCEPH | 138 | 23.7521515 | 21.8855219 | 25.7042254 | 3.81870347 | 0.27961485 | Chi-square |
| BIO_TROPO | 133 | 22.8915663 | 22.5589226 | 23.2394366 | 0.68051406 | 0.84526526 | Chi-square |
| J0_CILIAIRE | 130 | 22.3752151 | 22.5589226 | 22.1830986 | 0.37582397 | 0.91347608 | Chi-square |
| J0_CORNEEN | 107 | 18.4165232 | 18.1818182 | 18.6619718 | 0.48015365 | 0.88135358 | Chi-square |
| BIO_CALCIUM | 106 | 18.2444062 | 18.1818182 | 18.3098592 | 0.12804097 | 0.96813608 | Chi-square |
| V0_PLANCHE | 85 | 14.6299484 | 15.8249158 | 13.3802817 | 2.44463413 | 0.40458073 | Chi-square |
| BIO_GLYCEMI | 74 | 12.7366609 | 12.1212121 | 13.3802817 | 1.25906957 | 0.64907611 | Chi-square |
| BIO_PROTID | 70 | 12.0481928 | 11.4478114 | 12.6760563 | 1.22824489 | 0.64938082 | Chi-square |
| BIO_LACTAT | 57 | 9.81067126 | 10.1010101 | 9.50704225 | 0.59396785 | 0.80986957 | Chi-square |
| J0_BMI | 54 | 9.29432014 | 8.08080808 | 10.5633803 | 2.4825722 | 0.3029121 | Chi-square |
| BIO_TP | 53 | 9.1222031 | 7.74410774 | 10.5633803 | 2.81927254 | 0.2380806 | Chi-square |
| J0_TAILLE | 52 | 8.95008606 | 7.40740741 | 10.5633803 | 3.15597287 | 0.182835 | Chi-square |
| BIO_TEMP | 52 | 8.95008606 | 8.75420875 | 9.15492958 | 0.40072082 | 0.86568911 | Chi-square |
| J0_RYTHM | 50 | 8.60585198 | 6.73400673 | 10.5633803 | 3.82937355 | 0.09992683 | Chi-square |
| ECG_QTC | 50 | 8.60585198 | 8.41750842 | 8.8028169 | 0.38530848 | 0.86851932 | Chi-square |
| BIO_FIO2 | 49 | 8.43373494 | 9.42760943 | 7.3943662 | 2.03324323 | 0.37800484 | Chi-square |
| J0_ADRE_DOS | 48 | 8.2616179 | 7.07070707 | 9.50704225 | 2.43633518 | 0.28628931 | Chi-square |
| BIO_PACO2 | 41 | 7.05679862 | 8.75420875 | 5.28169014 | 3.47251861 | 0.1023148 | Chi-square |
| J0_VT | 40 | 6.88468158 | 7.07070707 | 6.69014085 | 0.38056623 | 0.85628626 | Chi-square |
| BIO_PAO2 | 40 | 6.88468158 | 8.41750842 | 5.28169014 | 3.13581828 | 0.1356281 | Chi-square |
| BIO_BICARB | 40 | 6.88468158 | 7.74410774 | 5.98591549 | 1.75819225 | 0.40276834 | Chi-square |
| BIO_PH | 39 | 6.71256454 | 8.08080808 | 5.28169014 | 2.79911794 | 0.17773344 | Chi-square |
| J0_TEMOIN_MASSE | 34 | 5.85197935 | 7.74410774 | 3.87323944 | 3.87086831 | 0.04692094 | Chi-square |
| BIO_UREE | 28 | 4.81927711 | 4.37710438 | 5.28169014 | 0.90458576 | 0.61082135 | Chi-square |
| ECG_ANOMALI | 27 | 4.64716007 | 4.04040404 | 5.28169014 | 1.2412861 | 0.47739767 | Chi-square |
| BIO_PLAQ | 26 | 4.47504303 | 4.37710438 | 4.57746479 | 0.20036041 | 0.90704809 | Chi-square |
| J0_PUPILD_REA | 25 | 4.30292599 | 4.71380471 | 3.87323944 | 0.84056528 | 0.61770737 | Chi-square |
| BIO_CREAT | 25 | 4.30292599 | 4.37710438 | 4.22535211 | 0.15175226 | 0.92820306 | Chi-square |
| BIO_POTAS | 24 | 4.13080895 | 3.7037037 | 4.57746479 | 0.87376109 | 0.59678072 | Chi-square |
| BIO_LEUCO | 23 | 3.95869191 | 3.7037037 | 4.22535211 | 0.52164841 | 0.74719056 | Chi-square |
| BIO_HEMO | 22 | 3.78657487 | 3.7037037 | 3.87323944 | 0.16953573 | 0.91477223 | Chi-square |
| J0_PUPILG_REA | 21 | 3.61445783 | 5.05050505 | 2.11267606 | 2.93782899 | 0.05789658 | Chi-square |
| BIO_SODIUM | 21 | 3.61445783 | 3.36700337 | 3.87323944 | 0.50623607 | 0.74382324 | Chi-square |
| J0_PUPILD | 19 | 3.27022375 | 3.7037037 | 2.81690141 | 0.8868023 | 0.54799404 | Chi-square |
| J0_PUPILG | 17 | 2.92598967 | 4.04040404 | 1.76056338 | 2.27984066 | 0.10311868 | Chi-square |
| V0_REFROIDI | 14 | 2.40963855 | 3.7037037 | 1.05633803 | 2.64736568 | 0.03751652 | Chi-square |
| J0_FR | 14 | 2.40963855 | 2.35690236 | 2.46478873 | 0.10788638 | 0.93244512 | Chi-square |
| J0_PEP | 9 | 1.54905336 | 1.34680135 | 1.76056338 | 0.41376203 | 0.7470861 | Fisher |
| J0_SPO2 | 8 | 1.37693632 | 2.02020202 | 0.70422535 | 1.31597667 | 0.28664891 | Fisher |
| J0_NOFLOW | 6 | 1.03270224 | 1.01010101 | 1.05633803 | 0.04623702 | 1 | Fisher |
| J0_FIO2 | 6 | 1.03270224 | 1.01010101 | 1.05633803 | 0.04623702 | 1 | Fisher |
| J0_TEMP | 5 | 0.8605852 | 0.33670034 | 1.4084507 | 1.07175037 | 0.20708218 | Fisher |
| J0_AGE | 4 | 0.68846816 | 0.67340067 | 0.70422535 | 0.03082468 | 1 | Fisher |
| J0_POIDS | 3 | 0.51635112 | 0.67340067 | 0.35211268 | 0.321288 | 1 | Fisher |
| J0_PAD | 2 | 0.34423408 | 0.33670034 | 0.35211268 | 0.01541234 | 1 | Fisher |
| J0_PAM | 2 | 0.34423408 | 0.67340067 | 0 | 0.67340067 | 0.49938869 | Fisher |
| J0_IGSII | 2 | 0.34423408 | 0 | 0.70422535 | 0.70422535 | 0.23850674 | Fisher |
| V0_CHOC_AV | 2 | 0.34423408 | 0 | 0.70422535 | 0.70422535 | 0.23850674 | Fisher |
| V0_CHOC_AP | 2 | 0.34423408 | 0 | 0.70422535 | 0.70422535 | 0.23850674 | Fisher |
| J0_PAS | 1 | 0.17211704 | 0.33670034 | 0 | 0.33670034 | 1 | Fisher |
| J0_FC | 1 | 0.17211704 | 0.33670034 | 0 | 0.33670034 | 1 | Fisher |
| J0_CAUSE2_ACR | 1 | 0.17211704 | 0 | 0.35211268 | 0.35211268 | 0.48881239 | Fisher |
| J0_TEMOIN | 1 | 0.17211704 | 0.33670034 | 0 | 0.33670034 | 1 | Fisher |
| J0_LOWFLOW | 1 | 0.17211704 | 0 | 0.35211268 | 0.35211268 | 0.48881239 | Fisher |
| J0_MCCABE | 0 | 0 | 0 | 0 | 0 |  | Failed |
| J0_KNAUS | 0 | 0 | 0 | 0 | 0 |  | Failed |
| J0_CHARLSON1 | 0 | 0 | 0 | 0 | 0 |  | Failed |
| J0_CHARLSON2 | 0 | 0 | 0 | 0 | 0 |  | Failed |
| J0_CHARLSON3 | 0 | 0 | 0 | 0 | 0 |  | Failed |
| J0_CHARLSON4 | 0 | 0 | 0 | 0 | 0 |  | Failed |
| J0_CHARLSON5 | 0 | 0 | 0 | 0 | 0 |  | Failed |
| J0_CHARLSON6 | 0 | 0 | 0 | 0 | 0 |  | Failed |
| J0_CHARLSON7 | 0 | 0 | 0 | 0 | 0 |  | Failed |
| J0_CHARLSON8 | 0 | 0 | 0 | 0 | 0 |  | Failed |
| J0_CHARLSON9 | 0 | 0 | 0 | 0 | 0 |  | Failed |
| J0_CHARLSON10 | 0 | 0 | 0 | 0 | 0 |  | Failed |
| J0_CHARLSON11 | 0 | 0 | 0 | 0 | 0 |  | Failed |
| J0_CHARLSON12 | 0 | 0 | 0 | 0 | 0 |  | Failed |
| J0_CHARLSON13 | 0 | 0 | 0 | 0 | 0 |  | Failed |
| J0_CHARLSON14 | 0 | 0 | 0 | 0 | 0 |  | Failed |
| V0_CHARLSON15 | 0 | 0 | 0 | 0 | 0 |  | Failed |
| V0_CHARLSON16 | 0 | 0 | 0 | 0 | 0 |  | Failed |
| V0_CHARLSON17 | 0 | 0 | 0 | 0 | 0 |  | Failed |
| V0_CHARLSON18B | 0 | 0 | 0 | 0 | 0 |  | Failed |
| V0_CHARLSON19 | 0 | 0 | 0 | 0 | 0 |  | Failed |
| J0_CHARLSON | 0 | 0 | 0 | 0 | 0 |  | Failed |
| J0_ATCD | 0 | 0 | 0 | 0 | 0 |  | Failed |
| J0_LIEU_ACR | 0 | 0 | 0 | 0 | 0 |  | Failed |
| J0_ADRE | 0 | 0 | 0 | 0 | 0 |  | Failed |
| J0_CORDA | 0 | 0 | 0 | 0 | 0 |  | Failed |
| J0_BICARB | 0 | 0 | 0 | 0 | 0 |  | Failed |
| V0_THROMBO | 0 | 0 | 0 | 0 | 0 |  | Failed |
| V0_CORO_ACR | 0 | 0 | 0 | 0 | 0 |  | Failed |
| V0_ANGIO_ACR | 0 | 0 | 0 | 0 | 0 |  | Failed |
| V0_BALLON | 0 | 0 | 0 | 0 | 0 |  | Failed |
| V0_ACR2 | 0 | 0 | 0 | 0 | 0 |  | Failed |
| J0_CURAR | 0 | 0 | 0 | 0 | 0 |  | Failed |
| J0_SEDATIF | 0 | 0 | 0 | 0 | 0 |  | Failed |
| J0_MORPHIN | 0 | 0 | 0 | 0 | 0 |  | Failed |
| J0_COAGUL | 0 | 0 | 0 | 0 | 0 |  | Failed |
| J0_AGREG | 0 | 0 | 0 | 0 | 0 |  | Failed |
| J0_ANTIBIO | 0 | 0 | 0 | 0 | 0 |  | Failed |
| J0_AMINE | 0 | 0 | 0 | 0 | 0 |  | Failed |
| ECG | 0 | 0 | 0 | 0 | 0 |  | Failed |
| SOFA_SC | 0 | 0 | 0 | 0 | 0 |  | Failed |
| SOFA_RESPIR | 0 | 0 | 0 | 0 | 0 |  | Failed |
| SOFA_CARDIO | 0 | 0 | 0 | 0 | 0 |  | Failed |
| SOFA_COAG | 0 | 0 | 0 | 0 | 0 |  | Failed |
| SOFA_NEURO | 0 | 0 | 0 | 0 | 0 |  | Failed |
| SOFA_FOIE | 0 | 0 | 0 | 0 | 0 |  | Failed |
| SOFA_RENAL | 0 | 0 | 0 | 0 | 0 |  | Failed |
| EI_EI | 0 | 0 | 0 | 0 | 0 |  | Failed |
| SOFA_SC1 | 0 | 0 | 0 | 0 | 0 |  | Failed |
| SEX | 0 | 0 | 0 | 0 | 0 |  | Failed |

**eTable 19** – Complete feature list and missing variable report for HYPERION dataset.

### S.13 STROBE Checklist

| **Item No.** | **Item** | **Recommendation** | **Location in Manuscript** |
| --- | --- | --- | --- |
| 1 | Title & Abstract | (a) Indicate the study's design with a commonly used term in the title or the abstract. (b) Provide in the abstract an informative and balanced summary of what was done and what was found. | (a) Title; (b) Abstract. |
| 2 | Background/rationale | Explain the scientific background and rationale for the investigation being reported. | Introduction. |
| 3 | Objectives | State specific objectives, including any prespecified hypotheses. | Introduction (final paragraph). |
| 4 | Study design | Present key elements of study design early in the paper. | Methods – Data sources. |
| 5 | Setting | Describe the setting, locations, and relevant dates, including periods of recruitment, exposure, follow-up, and data collection. | Methods – Data sources; eTable 1. |
| 6 | Participants | (a) Cohort study—give eligibility criteria, sources of data and methods of selection. (b) For matched studies, give matching criteria and number of exposed and unexposed. | (a) Methods – Patient Selection and TTM identification; eTable 1; eFigure 6. (b) Not applicable. |
| 7 | Variables | Clearly define all outcomes, exposures, predictors, potential confounders, and effect modifiers. Give diagnostic criteria, if applicable. | Methods – Predictive features, outcomes, missingness; eFigure 1; eTables 16-19; eTables 5-6. |
| 8 | Data sources/measurement | For each variable of interest, give sources of data and details of methods of assessment (measurement). Describe comparability of assessment methods if there is more than one group. | Methods – Predictive features, outcomes, missingness; eTables 16-19. |
| 9 | Bias | Describe any efforts to address potential sources of bias. | Methods – Causal machine learning framework (propensity diagnostics, eFigure 2); Methods - HTE assessment; Methods - Patient Selection and TTM identification (Figure 4 in main manuscript); Discussion - Limitations; eFigures 3-4. |
| 10 | Study size | Explain how the study size was arrived at. | Methods - Data sources; Discussion - Limitations (Statistical power for HTE detection); eTables 7-14. |
| 11 | Quantitative variables | Explain how quantitative variables were handled in the analyses. If applicable, describe which groupings were chosen and why. | Methods - Causal machine learning framework; eFigure 1. |
| 12 | Statistical methods | (a) Describe all statistical methods, including those used to control for confounding. (b) Describe any methods used to examine subgroups and interactions. (c) Explain how missing data were addressed. (d) If applicable, describe analytical methods taking account of sampling strategy. (e) Describe any sensitivity analyses. | (a) Methods - Causal machine learning framework and HTE assessment; eTable 2. (b) Methods - HTE assessment. (c) Methods - Predictive features, outcomes, missingness; eFigure 1. (d) Not applicable. (e) eTables 7-14; eTables 3-4; Discussion - Limitations. |
| 13 | Participants | (a) Report numbers of individuals at each stage of study. (b) Give reasons for non-participation at each stage. (c) Consider use of a flow diagram. | (a) Results - Study population; Table 1; eFigure 6. (b) eFigure 6. (c) eFigure 6. |
| 14 | Descriptive data | (a) Give characteristics of study participants. (b) Indicate number of participants with missing data for each variable of interest. | (a) Table 1. (b) eTables 16-19. |
| 15 | Outcome data | Report numbers of outcome events or summary measures over time. | Table 1; Results - Study population. |
| 16 | Main results | (a) Give unadjusted estimates and, if applicable, confounder-adjusted estimates. (b) Report category boundaries when continuous variables were categorized. (c) If relevant, consider translating estimates of relative risk into absolute risk for a meaningful time period. | (a) Table 2; Figure 2; Figure 3; eFigure 5; eTable 15. (b) Not applicable. (c) Not applicable. |
| 17 | Other analyses | Report other analyses done—e.g. analyses of subgroups and interactions, and sensitivity analyses. | Figure 3 and eTable 15 (GATES); eTables 5-6 (SHAP); eFigures 3-4 and eTables 3-4 (calibration); eTables 7-14 (power simulations). |
| 18 | Key results | Summarise key results with reference to study objectives. | Discussion - opening paragraph. |
| 19 | Limitations | Discuss limitations of the study, taking into account sources of potential bias or imprecision. | Discussion - Limitations. |
| 20 | Interpretation | Give a cautious overall interpretation of results considering objectives, limitations, multiplicity of analyses, and results from similar studies. | Discussion - Interpretation in context. |
| 21 | Generalisability | Discuss the generalisability (external validity) of the study results. | Discussion - Strengths; Discussion - Clinical implications. |
| 22 | Funding | Give the source of funding and the role of the funders for the present study and, if applicable, for the original study on which the present article is based. | No funding was received for this article. |

**eTable 20** – STROBE Checklist.
